# Risk-stratified monitoring for sulfasalazine toxicity: prognostic model development and validation

**DOI:** 10.1101/2023.12.15.23299947

**Authors:** A Abhishek, Matthew J Grainge, Tim Card, Hywel C Williams, Maarten W Taal, Guruprasad P Aithal, Christopher P Fox, Christian D Mallen, Matthew D Stevenson, Georgina Nakafero, Richard D Riley

**Affiliations:** Academic Rheumatology, School of Medicine, University of Nottingham, Nottingham NG5 1PB, UK; Lifespan and Population Health, School of Medicine, University of Nottingham, Nottingham NG5 1PB, UK; Centre for Kidney Research and Innovation, Translational Medical Sciences, School of Medicine, University of Nottingham, Derby DE22 3NE, UK; Nottingham Digestive Diseases Centre, Translational Medical Sciences, School of Medicine, University of Nottingham, Nottingham NG7 2UH, UK; Translational Medical Sciences, School of Medicine, University of Nottingham, Nottingham, UK; Primary Care Centre Versus Arthritis, School of Medicine, Keele University, Keele ST5 5BJ, UK; School of Health and Related Research, University of Sheffield, Sheffield S1 4DA, UK; Institute of Applied Health Research, College of Medical and Dental Sciences, University of Birmingham, Birmingham B15 2TT, UK

**Author notes:** **Corresponding author:** Prof. Abhishek Address for correspondence: A23, Academic Rheumatology, Clinical Sciences Building, The University of Nottingham, Nottingham NG5 1PB, UK. Contributed equally (Georgina Nakafero, Richard D Riley).

## Abstract

**Background:** Sulfasalazine induced cytopenia, nephrotoxicity, and hepatotoxicity is uncommon during long-term treatment. Some guidelines recommend three monthly monitoring blood-tests indefinitely while others recommend stopping monitoring after one year. To rationalise monitoring we developed and validated a prognostic model for clinically significant blood, liver, or kidney toxicity during established sulfasalazine treatment.

**Design:** Retrospective cohort study.

**Setting:** UK primary-care. Data from Clinical Practice Research Datalink Gold and Aurum formed independent development and validation cohorts.

**Participants:** Age ≥18 years, new diagnosis of an inflammatory condition and sulfasalazine prescription.

**Study period:** 01/01/2007 to 31/12/2019.

**Outcome:** Sulfasalazine discontinuation with abnormal monitoring blood-test result. *Analysis:* Patients were followed-up from six months after first primary-care prescription to the earliest of outcome, drug discontinuation, death, 5 years, or 31/12/2019.Penalised Cox regression was performed to develop the risk equation. Multiple imputation handled missing predictor data. Model performance was assessed in terms of calibration and discrimination.

**Results:** 8,936 participants were included in the development cohort (473 events, 23,299 person-years) and 5,203 participants were included in the validation cohort (280 events, 12,867 person-years).Nine candidate predictors were included. The optimism adjusted R^2^_D_ and Royston D statistic in the development data were 0.13 and 0.79 respectively. The calibration slope (95% confidence interval (CI)) and Royston D statistic (95% CI) in validation cohort was 1.19 (0.96-1.43) and 0.87 (0.67-1.07) respectively.

**Conclusion:** This prognostic model for sulfasalazine toxicity utilises readily available data and should be used to risk-stratify blood-test monitoring during established sulfasalazine treatment.<colcnt=1>

**Evidence before this study?:**

- Hepatic, haematological, and renal toxicity from sulfasalazine occurs uncommonly after the first-few months of treatment. Nevertheless, the manufacturers and some specialist societies e.g., the American College of Rheumatology recommend monitoring blood-tests at three monthly intervals during established treatment. Other guidelines e.g., from the British Society of Rheumatology recommend no monitoring after the first two years of treatment.
- It is not known whether hepatic, haematological, and renal toxicities due to sulfasalazine can be predicted and monitoring be risk-stratified.

**Added value of this study?:**

- This study developed a prognostic model that discriminated patients at varying risk of sulfasalazine toxicity during long-term treatment. It had excellent performance characteristics in an independent validation cohort.
- The model performed well across age-groups, and in people with rheumatoid arthritis and other inflammatory conditions.
- Any cytopenia or liver enzyme elevation prior to start of follow-up, chronic kidney disease stage-3, diabetes, methotrexate prescription, leflunomide prescription, and age were strong predictors of sulfasalazine toxicity.

**Implications of all the available evidence:**

- This prognostic model utilises information that can be easily ascertained during clinical visits. It can be used to inform decisions on the interval between monitoring blood-tests.
- The results of this study ought to be considered by national and international Rheumatology guideline writing groups to rationalise monitoring during long-term sulfasalazine treatment.

## Introduction

Sulfasalazine is commonly used in the treatment of inflammatory diseases such as rheumatoid arthritis (RA), psoriatic arthritis (PsA), axial spondylarthritis, reactive arthritis, and infrequently in the management of inflammatory bowel disease (IBD) (the latter is mostly treated with 5-aminosalicylates due to a better safety profile)^1–3^. Although effective, sulfasalazine can cause cytopenia and elevated liver enzymes typically in the first three to six months of treatment, although late onset toxicity is reported^4–16^. Sulfasalazine can also cause crystalluria and interstitial nephritis, and is not recommended in those with severe renal impairment^17^. Cautious use is recommended in those with mild to moderate renal impairment^17^.

There is considerable inconsistency in guidance on how to monitor patients on long-term sulfasalazine treatment for asymptomatic bone marrow, liver and/or renal toxicity. The British Society of Rheumatology (BSR) guidelines recommend two to four weekly blood-tests for full blood count (FBC), liver function test (LFT), urea electrolytes and creatinine (UE&C) for the first three months of treatment followed by three-monthly testing in the first year and no further monitoring blood-tests thereafter^18^. On the contrary, the American College of Rheumatology (ACR) guidelines recommend close monitoring for the first three months of treatment, followed by three-monthly blood-testing for FBC, UE&C, and LFT during the entire duration of treatment^19^. The summary of product characteristics for sulfasalazine recommends monitoring with FBC, LFT and UE&C at three monthly intervals during long-term treatment^20^. However, whether everyone needs a fixed monitoring schedule once established on sulfasalazine treatment, or whether monitoring can be risk-stratified during long-term treatment is not known.

To predict clinically significant laboratory abnormalities during established sulfasalazine treatment and to inform the frequency of testing, we have developed and validated a prognostic model for clinically significant myelotoxicity, hepatotoxicity and/or nephrotoxicity due to sulfasalazine.

## Methods

### Data source

Data from the Clinical Practice Research Datalink (CPRD) Aurum and Gold were used for model development and validation respectively ^21,22^. CPRD is an anonymised longitudinal database of electronic health records originated during clinical care in the National Health Service in the UK. With almost universal coverage of UK residents, participants that contributed data to the CPRD are representative of the UK population^21^. The CPRD includes information on demographic details, lifestyle factors (e.g., smoking, alcohol intake), diagnoses, results of blood-tests, and details of primary-care prescriptions. CPRD Gold and Aurum complement each other in terms of coverage of general-practices due to their use of different software for data capture. Some general practices that have contributed data to both databases are identifiable using a bridging file provided by the CPRD.

### Approvals

Independent Scientific Advisory Committee of the MHRA (Reference: 19_275R, 20_000236R).

### Study design

Retrospective cohort study.

### Study period

1^st^ January 2007 to 31^st^ December 2019.

### Study population

Participants aged 18 years or older with a new diagnosis of inflammatory disease (e.g., RA, axial spondyloarthritis, PsA, IBD etc.) and prescribed sulfasalazine by their GP for ≥six months were eligible. Patients were required to have ≥1-year disease-free registration in their current general practice to be classified as having a new diagnosis^23^.Additionally, patients were required to have received their first sulfasalazine prescription either after the first record of inflammatory disease in the CPRD or in the 90-days preceding. This 90-day period was allowed because recording of diagnosis may lag prescriptions. These two requirements minimised the chance of patients on long-term sulfasalazine treatment appearing as new users of sulfasalazine when they moved to a different general practice. Patients with chronic liver disease, haematological disease, and chronic kidney disease (CKD) stage 4 or 5 prior to cohort entry were excluded as described in a previous manuscript^24^.

### Sulfasalazine prescriptions

In the UK, sulfasalazine initiation and dose-escalation occur in hospital out-patient clinics. During this period prescriptions are issued by the hospital specialists. They also organise monitoring blood-tests and acts on any abnormalities. Once a patient is established on treatment, typically approximately six months after initiating on treatment, the responsibility for prescribing and monitoring, including with periodic blood-tests is handed to the patients’ general practitioner (GP) as per the NHS shared-care protocols. During shared-care monitoring, the GP seeks advice from the hospital specialist if there are side-effects including abnormal blood-test results, and treatment changes are directed by the specialist.

### Start of follow-up

Patients were followed-up from 180 days after their first primary-care sulfasalazine prescription until the earliest of outcome, death, transfer out of practice, 90-days prescription gap, last data collection from practice, 31/12/2019 or five-years.

### Outcome

Sulfasalazine-toxicity associated drug discontinuation was the outcome of interest. This was defined as a prescription gap of ≥90 days with either an abnormal blood-test result or a diagnostic code for abnormal blood-test result within ±60 days of the last prescription date^25^.The blood tests were considered abnormal if any of the following were present: total leucocyte count <3.5×10^9^/L, neutrophil count <1.6×10^9^/L, platelet count <140×10^9^/L, alanine transaminase and/or aspartate transaminase >100 IU/mL, and decline in kidney function, defined as either progression of chronic kidney disease based on medical codes recorded by the GP, or >26 μmol/L increase in creatinine concentration, the threshold for consideration of acute kidney injury^18,26^.In a previous validation study on methotrexate discontinuation, only 5.4% of abnormal blood-test results in this time-window were potentially explained by an alternate illness^25^.

A random sample of sulfasalazine discontinuations with abnormal blood test results was drawn. Data for all diagnostic codes entered during primary-care consultations within ±60 days of the abnormal blood test result were extracted. A.A. screened the list to identify outcomes that could potentially be explained by an alternative condition or its treatment.

### Predictors

These were selected by the clinical members of the study-team based on their clinical expertise and knowledge of the published literature. Age, sex, body mass index (BMI), alcohol intake, and diabetes were included as they associate with drug induced liver injury (DILI) ^27,28^. Individual inflammatory diseases were considered separately because sulfasalazine toxicity is reported to be less common in people with inflammatory bowel disease than in those with RA^3^. CKD stage-3 was included as it reduces sulfasalazine clearance^29^. Statins, carbamazepine, valproate, and paracetamol were included as their use is associated with sulfasalazine toxicity as per the British National Formulary. Methotrexate, leflunomide, thiopurines were included as they can cause cytopenia, elevated liver enzymes and acute kidney injury (AKI).Either cytopenia (neutrophil count <2 x 10^9^/l, total leucocyte count <4 x 10^9^/l, or platelet count <150 10^9^/l) or elevated transaminase (ALT and/or AST >35 IU/l) during the first six months of primary-care prescription were included as they predicted cytopenia and/or transaminitis in other studies ^30,31^.

The latest record of demographic and lifestyle factors, diseases recorded within two years prior to start of follow-up, and latest primary-care prescriptions within six-month prior to start of follow-up were used to define predictors except for CKD stage-3 that was defined using both GP records and/or eGFR 30-59 ml/min. GPs typically review patients with long-term conditions annually. A two-year look-back was utilised to minimise the risk of missing data from those that did not attend in a year.

### Patient and public involvement (PPI)

PPI members were involved in selecting and prioritising the research question. They advised to use readily available datasets for the study rather than conduct an expensive and time-consuming clinical trial.

### Sample size

In a previously published cohort of 1,321 RA patients, 85 stopped sulfasalazine with neutropenia, thrombocytopenia, or elevated liver enzymes during a mean follow-up of 2.39 years^16^. Assuming a similar incidence of treatment discontinuation for model development, the minimum sample size needed to minimise model overfitting (a target shrinkage factor of 0.9) and ensure precise estimation of overall risk was 1,748 participants (113 outcomes) based on a maximum of 25 parameters, Cox-Snell R^2^ value of 0.12, outcome rate of 0.027/person-year^16^, a 5-year time horizon, and a mean follow-up period of 2.39 years using the formulae of Riley et al.^32^. The sample size for external model validation was much larger than the typically recommended minimum sample size of 200 events ^33^.

### Statistical analysis

Multiple imputation handled missing data on BMI, alcohol intake, and sulfasalazine dose using chained equations ^34^. We carried out 10 imputations in the development dataset and five imputations in the validation dataset - a pragmatic approach considering the larger size of CPRD Aurum. The imputation model included all candidate predictors, Nelson-Aalen cumulative hazard function and outcome variable. The data analysis was undertaken using the Stata command “mi estimate” in a combined dataset that included all imputations.

### Model development

Fraction polynomial regression (first degree) analysis was used to model non-linear risk relationships with continuous predictors, but these were not better than the linear terms (p > 0.05), hence were not transformed. All 12 candidate predictors (19 parameters) were included in the Cox model and coefficients of each parameter estimated and combined using Rubin’s rule across the imputed datasets. The risk equation for predicting an individual’s risk of sulfasalazine discontinuation with abnormal blood-test results by five-years follow-up was formulated in the development data. The baseline survival function at *t=5 years*, a non-parametric estimate of survival function when all predictor values are set to zero, which is equivalent to the Kaplan-Meier product-limit estimate, was estimated along with the estimated regression coefficients (β) and the individual’s predictor values (X).This led to the equation for the predicted absolute risk over time ^35^:

Predicted risk of sulfasalazine-toxicity associated drug discontinuation at 5-years =1 – S_0_(t_=5_)^exp(Xβ)^ where S_0_(t_=5_) is the baseline survival function at 5-years of follow-up and βX is the linear predictor, β_1_x_1_+ β_2_x_2_+ … + β_p_x_p_.

### Model internal validation and shrinkage

The performance of the model in terms of calibration (where 1.00 is the ideal) was assessed by plotting agreement between predicted and observed outcomes. Internal validation was performed to correct performance estimates for optimism due to overfitting by bootstrapping with replacement 500 samples of the development data. The full model was fitted in each bootstrap sample and then its performance was quantified in the bootstrap sample (apparent performance) and the original sample (test model performance), and the optimism calculated (difference in test performance and apparent performance).A uniform shrinkage factor was estimated as the average of calibration slopes from the bootstrap samples. This process was repeated for all 10 imputed datasets, and the final uniform shrinkage calculated by averaging across the estimated shrinkage estimates from each imputation. Optimism-adjusted estimates of performance for the original model were then calculated, as the original apparent performance minus the optimism.

To account for overfitting during model development process, the original β coefficients were multiplied by the final uniform shrinkage factor and the baseline hazards re-estimated conditional on the shrunken β coefficients to ensure that overall calibration was maintained, producing a final model. The D statistic, a measure of discrimination, interpreted as a log hazard ratio (HR), the exponential of which gives the HR comparing two groups defined by above/below the median of the linear predictor was calculated ^36,37^.R^2^, a measure of variation explained by the model was calculated.

### Model external validation

External validation of the final model was performed using data from CPRD Gold. The final developed model equation was applied to the validation dataset, and calibration and discrimination were examined using the same measures as above ^36,37^.Calibration of 5-year risks was examined by plotting agreement between estimated risk from the model and observed outcome risks. In the calibration plot, predicted and observed risks were divided into 10 equally sized groups. Additionally, pseudo-observations were used to construct smooth calibration curves across all individuals via a running non-parametric smoother. Separate graphs were plotted for each imputation of the validation cohort and an example of one plot is shown in the results. Subgroup analyses considered age-group and inflammatory disease type (RA vs. others). Stata-MP version 16 was used for all statistical analyses. This study was reported in line with the transparent reporting of a multivariate prediction model for individual prediction or diagnosis guidelines ^38^.

## Results

### Study participants

Data for 8,936 and 5,203 participants contributing 23,299 and 12,867 person-years follow-up were included in the derivation and validation cohorts, respectively (Supplementary figure S1 and S2). Most participants in both cohorts were diagnosed with RA, were female, and had similar prevalence of lifestyle factors, comorbidities and drug treatments (Table 1). Nine candidate predictors (21 parameters) were included in the model (Table 2).

**Table 1:** Distribution of candidate predictors in development and validation cohorts

| Predictor<sup>1</sup> | Development cohort<br>(CPRD Aurum)<br>n=8,936 | Validation cohort<br>(CPRD Gold)<br>n=5,203 |
| --- | --- | --- |
| Age, mean (SD) year | 55.3 (14.8) | 55.5 (14.8) |
| Female sex | 5,535 (61.9) | 3,240 (62.3) |
| Body Mass Index |  |  |
| <18.5 kg/m<sup>2</sup> | 138 (1.5) | 88 (1.7) |
| 18.5-24.9 kg/m<sup>2</sup> | 2,441 (27.3) | 1,428 (27.5) |
| 25.0-29.9 kg/m<sup>2</sup> | 2,840 (31.8) | 1,678 (32.3) |
| ≥30 kg/m<sup>2</sup> | 2,714 (30.4) | 1,626 (31.3) |
| Missing | 803 (9.0) | 383 (7.4) |
| Alcohol use |  |  |
| Non-user | 1,705 (19.1) | 805 (15.5) |
| Low (1-14 units/week) | 3,854 (43.1) | 2,859 (55.0) |
| Moderate (15-21 units/week) | 535 (6.0) | 251 (4.8) |
| Hazardous (>21 units/week) | 667 (7.5) | 273 (5.3) |
| Ex-user | 996 (11.2) | 359 (6.9) |
| Missing | 1,179 (13.2) | 656 (12.6) |
| Inflammatory conditions |  |  |
| Rheumatoid arthritis | 6,945 (77.7) | 4,067 (78.2) |
| Psoriatic arthritis | 1,354 (15.2) | 773 (14.9) |
| Inflammatory bowel disease | 319 (3.6) | 173 (3.3) |
| Ankylosing spondylitis/reactive arthritis | 318 (3.6) | 190 (3.7) |
| Comorbidities |  |  |
| Diabetes | 982 (11.0) | 519 (10.0) |
| Chronic kidney disease stage-3 | 613 (6.9) | 333 (6.4) |
| Immunosuppressive drugs |  |  |
| Methotrexate | 2,999 (33.6) | 1,785 (34.3) |
| Leflunomide | 109 (1.2) | 78 (1.5) |
| Azathioprine/mercaptopurine | 73 (0.8) | 41 (0.8) |
| Other drugs |  |  |
| Statins | 2,088 (23.4) | 1,130 (21.7) |
| Carbamazepine/valproate | 103 (1.2) | 37 (0.7) |
| Paracetamol | 1,445 (16.2) | 884 (17.0) |
| At least mild cytopenia or liver enzyme elevation in six-months preceding start of follow-up | 1,264 (14.2) | 753 (14.5) |

<sup>1</sup>Values are numbers (percentage) unless stated otherwise.

**Table 2:** Final model hazard ratios and β-coefficients

| c | Adjusted HR<br>(95% CI) | Coefficients |
| --- | --- | --- |
| Age, mean (SD) year | 1.01 (1.00,1.02) | .0076439 |
| Female sex | 1.08 (0.88, 1.31) | .0741336 |
| Body Mass Index | 0.98 (0.97,1.00) | -.0168035 |
| Alcohol use |  |  |
| Non-user | Reference |  |
| Low (1-14 units/week) | 1.02 (0.80, 1.29) | .0182851 |
| Moderate (15-21 units/week) | 0.64 (0.38,1.06) | -.4507257 |
| Hazardous (>21 units/week) | 0.87 (0.58,1.33) | -.133557 |
| Ex-user | 0.94 (0.67,1.32) | -.0651469 |
| Inflammatory conditions |  |  |
| Rheumatoid arthritis | Reference |  |
| Psoriatic arthritis | 1.03 (0.78,1.36) | .0316689 |
| Inflammatory bowel disease | 0.74 (0.38,1.44) | -.305206 |
| Ankylosing spondylitis/reactive arthritis | 1.25 (0.74,2.12) | .2214547 |
| Comorbidities |  |  |
| Diabetes | 1.34 (1.01, 1.78) | .2909969 |
| Chronic kidney disease stage-3 | 1.96 (1.47,2.62) | .671859 |
| Immunosuppressive drugs |  |  |
| Methotrexate | 1.39 (1.15,1.68) | .3315573 |
| Leflunomide | 2.05 (1.09,3.86) | .7164324 |
| Azathioprine/mercaptopurine | 1.24 (0.37,4.17) | .2189764 |
| Other drugs |  |  |
| Statins | 0.98 (0.78,1.24) | -.0181917 |
| Carbamazepine/valproate | 0.74 (0.28,2.00) | -.2949835 |
| Paracetamol | 1.14 (0.90,1.43) | .1272515 |
| **Blood-test abnormalities** |  |  |
| At least mild cytopenia or liver enzyme elevation in six-months preceding start of follow-up | 2.80 (2.29,3.42) | 1.029245 |

<sup>1</sup>HR: hazard ratio, CI: confidence interval. The reported values are before shrinkage.

### Model development

In the derivation dataset, 473 outcome events occurred during the follow-up period at a rate (95% CI) of 20.30 (18.55 - 22.22) per 1,000 person-years. Of these, 256, 131, and 113 patients respectively stopped treatment due to cytopenia, renal function decline, and elevated liver enzymes. Outcome validation exercise in 178 outcomes revealed that only 4.5% outcomes (n=8) could potentially be explained by another contemporaneous illness or its treatments, with a positive predictive value of 95.5% (Table S1).

These events occurred throughout the 5-year follow-up period when the entire cohort was considered (Figure S3) and when patients co-prescribed either methotrexate or leflunomide or thiopurine with sulfasalazine were excluded (Figure S4). CKD-stage 3, diabetes (either type 1 or 2), co-prescription of methotrexate, co-prescription of leflunomide, and either cytopenia or elevated liver enzymes during first six months of sulfasalazine prescription were strong predictors of drug discontinuation with adjusted HR hazard ratio (95% CI) 1.96 (1.47-2.62), 1.34 (1.01-1.78), 1.39 (1.15-1.68), 2.05 (1.09-3.86) and 2.80 (2.29-3.42) respectively (Table 2). From the bootstrap, a uniform shrinkage factor of 0.84 was obtained and used to shrink predictor coefficients in the final model for optimism and after re-estimation, the final model’s cumulative baseline survival function (S_0_) was 0.940 at 5-years of follow-up (Box 1).

### Model performance in the development cohort

As expected, the calibration slope (95% CI) in the development data was1.00 (95% CI 0.85-1.15). Calibration plot of the final (i.e., after shrinkage) model at 5-years showed that the average model predictions matched the average observed outcome probabilities across 10 groups of patients, with confidence intervals overlapping the 45-degree line (perfect prediction line) (Figure 1). As most patients had a low risk of outcome (Figure S5), most of the deciles clustered at the bottom left of the calibration plot (Figure S6). The smoothed calibration curve at 5-years showed alignment of observed risk to the predicted risk with wide confidence intervals at high-risk probabilities (Figure 1). The Royston *D* statistic was 0.91 (95% CI 0.77 – 1.05), corresponding to a HR (95% CI) of 2.48 (2.16-2.86) comparing the risk of participants who were above the median of linear predictor to that below the median. The optimism adjusted Royston *D* statistic was 0.79, corresponding to a HR of 2.20 (Table 3).

**Figure 1:**
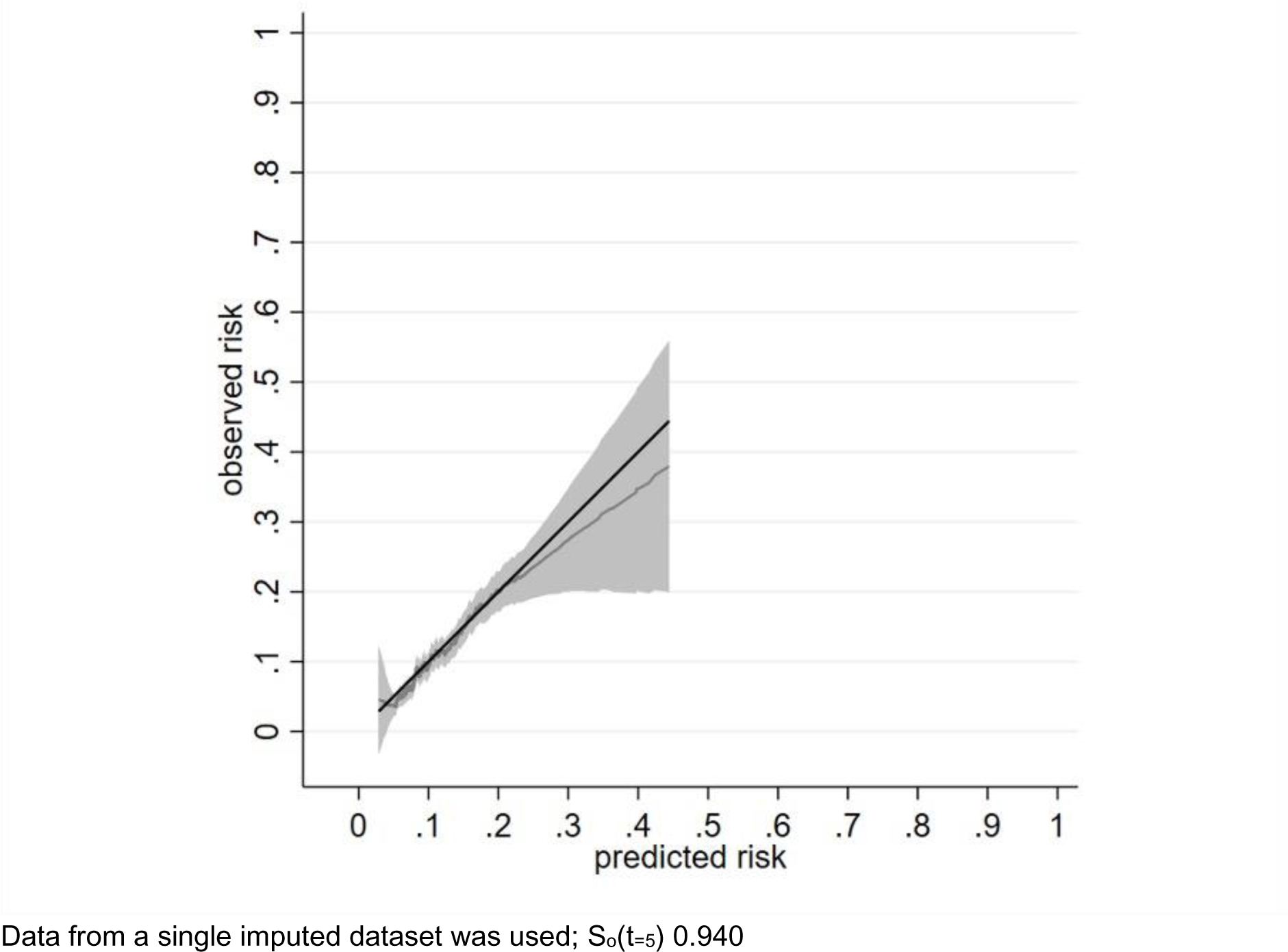
Calibration of a prognostic model for SSZ discontinuation with abnormal monitoring blood-test results at 5 years in the development cohort. Data from a single imputed dataset was used; So(t=5) 0.940

**Table 3:** Model diagnostics

| Measure | Apparent performance* | Test performance<sup>§</sup> | Average optimism<sup>¥</sup> | Optimism corrected performance<sup>†</sup> | External validation (CPRD Aurum)<sup>‡</sup> |
| --- | --- | --- | --- | --- | --- |
| Overall calibration slope | 1.00<br>(0.85,1.15) | 0.84<br>(0.70,0.98) | 0.16 | 0.84<br>(0.69,0.99) | 1.19<br>(0.96,1.43) |
| R<sup>2</sup><sub>D</sub> | 0.17<br>(0.12,0.21) | 0.15<br>(0.11,0.19) | 0.04 | 0.13<br>(0.08,0.17) | 0.15<br>(0.10,0.21) |
| Royston D statistic | 0.91<br>(0.77,1.05) | 0.85<br>(0.72,0.99) | 0.12 | 0.79<br>(0.65,0.93) | 0.87<br>(0.67,1.07) |

\*Refers to performance (95% CI) estimated directly from the data that was used to develop the model.
<sup>§</sup> Determined by executing full model in each bootstrap sample (500 samples with replacement), calculating bootstrap performance, and applying same model in original sample.
<sup>¥</sup> Average difference between model performance in bootstrap data and test performance in original dataset
<sup>†</sup> Subtracting average optimism from apparent performance.
<sup>‡</sup> Penalised model was externally validated (Penalised calibration slope:1.19; 95% CI 1.01, 1.37)
CPRD: Clinical Practice Research Datalink

### Model performance in the validation cohort

There were 280 outcomes at a rate (95% CI) of 21.76 (19.36-24.47)/1000 person-years in the validation cohort. The calibration slope (95% CI) across the 5-year follow-up period was 1.19 (0.96-1.43) (Figure 2). The calibration plot showed reasonable correspondence between observed and predicted risk at 5-years across the tenths of risk (Figure S7). Most of the deciles clustered at the bottom left of the calibration plot due to a low risk of outcome for most patients (Figures S7, S8). When individual risks were plotted, the smoothed calibration curve showed alignment of the predicted risk to the observed risk at low risk and wide confidence intervals overlapping the perfect prediction line at high-risk probabilities (Figure 2). Model performance was also tested at years 1, 2, 3 and 4 (Figure S9-S12) and showed a similar pattern except for over-prediction of risk at 1 year. The Royston D statistic in the validation data was 0.87 (0.67,1.07), corresponding to a HR (95% CI) of 2.39 (1.95-2.92). Model discrimination in the derivation and validation data was broadly similar (Table 3). The model performed well in those younger or older than 60 years, in those with RA or other conditions (Figure S13, S14).

**Figure 2:**
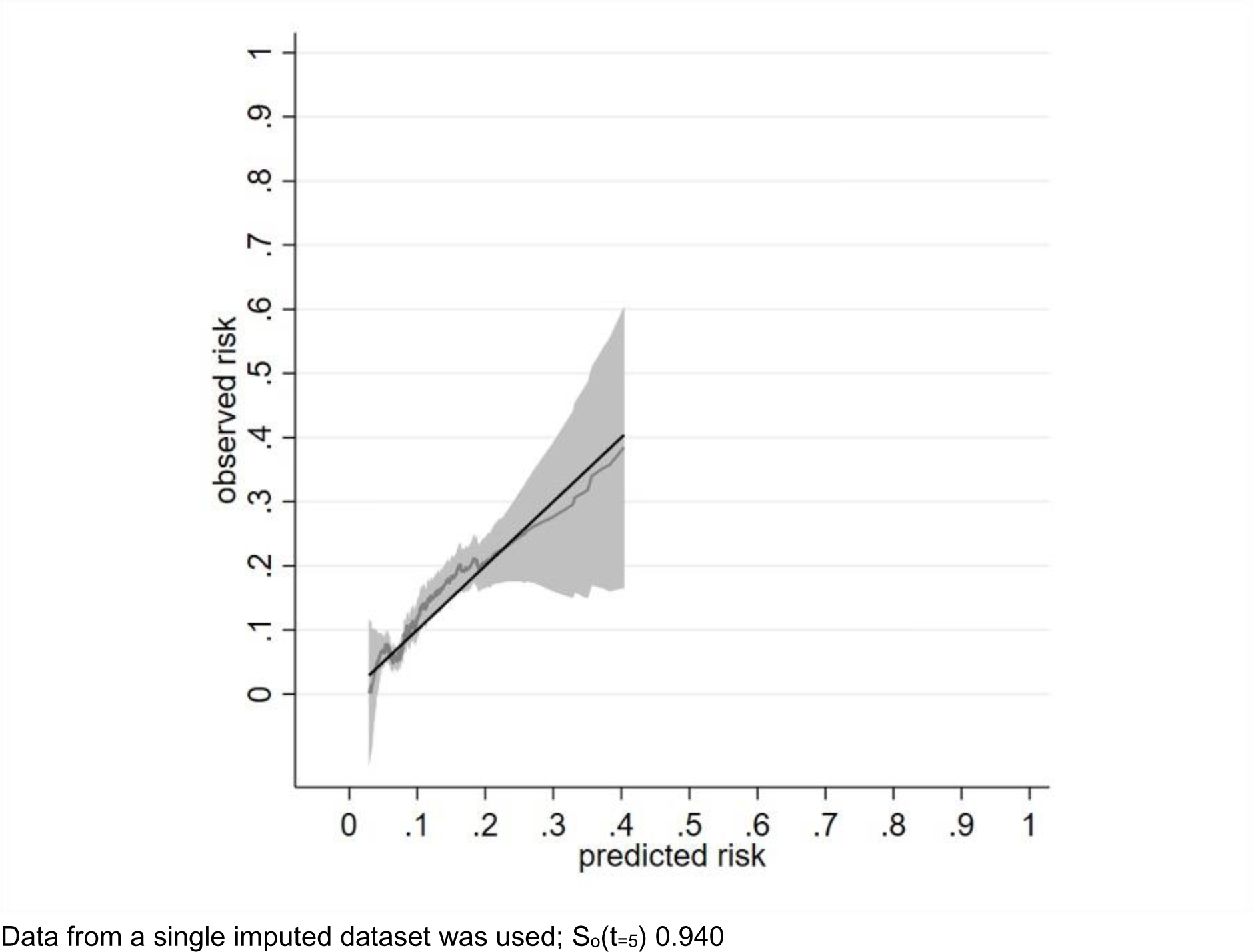
Calibration of a prognostic model for SSZ discontinuation with abnormal monitoring blood-test results at 5 years in the validation cohort. Data from a single imputed dataset was used; So(t=5) 0.940

Worked examples: Ten anonymised patient profiles, one from the middle of each of the 10 groups defined by deciles of predicted risk were selected from the development cohort, the higher the decile group the higher the risk, and the risk equation was applied to each. The cumulative probability of outcome over five years ranged from 5.3% in the middle of the first group to 9.3% in the middle of the seventh group, and 19.0% in the middle of the 10th group (Table S2).

## Discussion

We have developed and externally validated a prognostic model for sulfasalazine discontinuation due to abnormal blood-test results. To the best of our knowledge this is the first such risk-prediction model. It performed well in predicting outcomes by five years and in clinically relevant subgroups defined by age and inflammatory condition. Previous studies have variably reported NAT-2 acetylator status to be associated with sulfasalazine toxicity^15,39^. However, these studies evaluated all side-effects and did not separately assess either myelotoxicity, hepatotoxicity, or nephrotoxicity as evaluated in the current study.

Our findings suggest that a one size fits all approach to monitoring for blood, liver, or renal toxicity using three monthly blood-tests during long-term sulfasalazine treatment as recommended in the SmPC and the ACR guidelines, and not monitoring for these after the first year of treatment as recommended in the BSR guidelines are both inappropriate because there is a large interindividual variation in the risk of developing these side-effects. The large variation in risk implies that it may be reasonable to not monitor some patients after the first year of sulfasalazine treatment, while others at higher risk of side-effects are monitored frequently e.g., three-monthly. It is important to realise that DILI can be idiosyncratic and annual testing is unlikely to detect them early enough to improve patient outcome. It is beyond our remit to propose threshold at which the frequency of monitoring blood-tests should be altered. These decisions are best taken by guideline writing groups. Thus, our findings ought to be considered by guideline writing groups.

It is important that the results of this study are not used to risk-stratify monitoring in patients newly started on sulfasalazine because our prognosis model used data from patients prescribed sulfasalazine by their GP for six months after initiating treatment and dose-escalation in a hospital outpatient. It typically takes three to six months to stabilise a patients’ sulfasalazine dose before prescription and monitoring is handed over to the GP. In healthcare systems where such shared care arrangements do not exist, this strategy may be applied after one year of sulfasalazine treatment. Although generally perceived to be safe, sulfasalazine use carries a risk of myelotoxicity and hepatotoxicity comparable to that observed with methotrexate in people with RA^40^.

CKD stage-3, diabetes, and concomitant methotrexate or leflunomide therapy were strong independent predictors of sulfasalazine discontinuation with abnormal monitoring blood-test results in this study. These associations may be due to reduced sulfasalazine clearance in CKD and DILI being associated with diabetes^41^. Abnormal blood-test results during the first six-months of therapy were strong independent predictors of discontinuing sulfasalazine with abnormal monitoring blood-test results, like findings for methotrexate and leflunomide^24,42^. Elevated liver enzymes and cytopenia before starting treatment have previously been associated with abnormal blood-test results in patients treated with methotrexate and biologics respectively^43–49^.

There are several strengths of this study. First, we used a large real-world and nationally representative dataset for model development and a similar independent dataset for external validation. Second, the study population included patients with a range of diseases and the results have broad generalisability. Third, the prognostic factors were selected by an expert multidisciplinary team based on clinical experience. Fourth, our outcome required the abnormal blood-test result to be associated with sulfasalazine discontinuation, thus, allowing the model to predict clinically relevant outcomes. Fifth, the prognostic model is easy to use in practice, and can be easily built into GP electronic health records.

However, several limitations of this study ought to be considered. First, we did not have access to the date when the patient was first prescribed sulfasalazine in the hospital clinic. Second, we did not have data on concurrent use of biologics as these are hospital prescribed. However, there is no evidence to suggest that biologics increase sulfasalazine toxicity. Third, we did not have data on disease activity as these are not recorded in the CPRD. Fourth, the abnormal blood test could be due to a different illness and not due to sulfasalazine. However, in our previous validation studies on methotrexate, only 5.4% of abnormal blood-test results could be explained by an alternative illness ^25^. Fifth, although the external validation dataset was distinct from the model development dataset, it also originated from UK general practice. We recommend therefore that our model be validated in a dataset from another country. Sixth, there were 31 (0.3%) patients in the highest three risk groups defined according to tenths of risk, resulting in uncertainty regarding predictors for these groups. Seventh, we did not perform competing risk regression. However, this does not limit the validity of our findings as there were few deaths (28 [0.3%]) in the derivation cohort and 8 (0.2%) deaths in validation cohort up to 5-year follow-up period.

In conclusion, we have developed and externally validated a prognostic model for sulfasalazine discontinuation with abnormal monitoring blood-test results. These findings need to be considered by national and international specialist societies’ guideline writing groups to decide upon risk-stratified frequency of monitoring blood-tests during long-term sulfasalazine treatment.

## Ethics statements

### Ethical approval

Not required as this study is based on secondary data analysis.

## Footnotes

## Contributor and guarantor information

GN, MJG, HCW, TC, MWT, GPA, CPF, CDM, MDS, RDR, and AA designed the study.GN analysed the data supervised by MJG, RDR and AA.GN, MJG, HCW, TC, MWT, GPA, CPF, CDM, MDS, RDR, and AA interpreted the data.AA drafted the manuscript.All authors critically evaluated and revised the manuscript.The corresponding author attests that all listed authors meet authorship criteria and that no others meeting the criteria have been omitted.AA is the guarantor.

### Box 1

Equation to predict the risk of sulfasalazine discontinuation after six months of primary care prescription and within the next 5-years.

Risk score = 1 − 0.940 ^exp(0.84βX)^, where βX=(.0076439 * Age in years at first primary-care prescription + .0741336*female-sex - .0168035*BMI + .0182851*low alcohol intake - .4507257*moderate alcohol intake - .1335573*hazardous alcohol intake - .0651469*Ex-alcohol intake + .0316689*Psoriasis - .305206*IBD + .2214547*ankylosing spondylitis/reactive arthritis + .2909969*diabetes + .671859*CKD + .3315573*MTX + .7164324* LEF + .2189764*AZA or 6-MP - .0181917*statins - .2949835 *Carbamazepine/valproate + .1272515*paracetamol + 1.029245* At-least mild cytopenia or liver enzyme elevation within six-months of primary care AZA/6-MP prescription.

All variables are code 0, and 1 if absent or present respectively, except for BMI and age that were continuous variables.0.940 is the baseline survival function at 5-years, 0.84 is the shrinkage factor and the other numbers are the estimated regression coefficients for the predictors, which indicate their mutually adjusted relative contribution to the outcome risk.

## Data Availability

Data used in the study are from the Clinical Practice Research Datalink. Study protocol is available from www.cprd.com.

## References

1. Ogdie A, Coates LC, Gladman DD.Treatment guidelines in psoriatic arthritis.Rheumatology (Oxford) 2020;59(Suppl 1):i37–i46.doi: 10.1093/rheumatology/kez383 [published Online First: 2020/03/12]

2. Singh JA.Treatment Guidelines in Rheumatoid Arthritis.Rheum Dis Clin North Am 2022;48(3):679–89.doi: 10.1016/j.rdc.2022.03.005 [published Online First: 2022/08/12]

3. Ransford RA, Langman MJ.Sulphasalazine and mesalazine: serious adverse reactions re-evaluated on the basis of suspected adverse reaction reports to the Committee on Safety of Medicines.Gut 2002;51(4):536–9.doi: 10.1136/gut.51.4.536 [published Online First: 2002/09/18]

4. Farr M, Tunn EJ, Symmons DP, et al.Sulphasalazine in rheumatoid arthritis: haematological problems and changes in haematological indices associated with therapy.Br J Rheumatol 1989;28(2):134–8.doi: 10.1093/rheumatology/28.2.134 [published Online First: 1989/04/01]

5. McConkey B. Ten Years of Sulphasalazine Use in Rheumatoid Arthritis.Rheumatology 1989;28(2):175–76.doi: 10.1093/rheumatology/28.2.175

6. van Riel PLCM, van Gestel AM, van de Putte LBA.Long-Term Usage and Side-Effect Profile of Sulphasalazine in Rheumatoid Arthritis.Rheumatology 1995;XXXIV(suppl_4):40-42.doi: 10.1093/rheumatology/XXXIV.suppl_4.40

7. de Abajo FJ, Montero D, Madurga M, et al.Acute and clinically relevant drug-induced liver injury: a population based case-control study.Br J Clin Pharmacol 2004;58(1):71–80.doi: 10.1111/j.1365-2125.2004.02133.x [published Online First: 2004/06/23]

8. McConkey B, Amos RS, Durham S, et al.Sulphasalazine in rheumatoid arthritis.Br Med J 1980;280(6212):442-4.doi: 10.1136/bmj.280.6212.442 [published Online First: 1980/02/16]

9. Farr M, Scott DGI, Bacon PA.Side Effect Profile of 200 Patients with Inflammatory Arthritides Treated with Sulphasalazine.Drugs 1986;32(1):49–53.doi: 10.2165/00003495-198600321-00010

10. Amos RS, Pullar T, Bax DE, et al.Sulphasalazine for rheumatoid arthritis: toxicity in 774 patients monitored for one to 11 years.Br Med J (Clin Res Ed) 1986;293(6544):420-3.doi: 10.1136/bmj.293.6544.420 [published Online First: 1986/08/16]

11. MacGilchrist AJ, Hunter JA.Sulphasalazine hepatotoxicity: lack of a hypersensitivity response.Ann Rheum Dis 1986;45(11):967–8.doi: 10.1136/ard.45.11.967 [published Online First: 1986/11/01]

12. Marabani M, Madhok R, Capell HA, et al.Leucopenia during sulphasalazine treatment for rheumatoid arthritis.Ann Rheum Dis 1989;48(6):505–7.doi: 10.1136/ard.48.6.505 [published Online First: 1989/06/01]

13. Jobanputra P, Amarasena R, Maggs F, et al.Hepatotoxicity associated with sulfasalazine in inflammatory arthritis: A case series from a local surveillance of serious adverse events.BMC Musculoskelet Disord 2008;9:48.doi: 10.1186/1471-2474-9-48 [published Online First: 2008/04/15]

14. Keisu M, Ekman E.Sulfasalazine associated agranulocytosis in Sweden 1972– 1989.European Journal of Clinical Pharmacology 1992;43(3):215–18.doi: 10.1007/BF02333012

15. Wiese MD, Alotaibi N, O’Doherty C, et al.Pharmacogenomics of NAT2 and ABCG2 influence the toxicity and efficacy of sulphasalazine containing DMARD regimens in early rheumatoid arthritis.The Pharmacogenomics Journal 2014;14(4):350–55.doi: 10.1038/tpj.2013.45

16. Grove ML, Hassell AB, Hay EM, et al.Adverse reactions to disease-modifying anti-rheumatic drugs in clinical practice.Qjm 2001;94(6):309–19.doi: 10.1093/qjmed/94.6.309 [published Online First: 2001/06/08]

17. Committee JF.British National Formulary.[85].London: BMJ Group and Pharmaceutical Press; [2023].

18. Ledingham J, Gullick N, Irving K, et al.BSR and BHPR guideline for the prescription and monitoring of non-biologic disease-modifying anti-rheumatic drugs.Rheumatology (Oxford) 2017;56(6):865–68.doi: 10.1093/rheumatology/kew479 [published Online First: 2017/03/25]

19. Singh JA, Saag KG, Bridges SL, Jr., et al.2015 American College of Rheumatology Guideline for the Treatment of Rheumatoid Arthritis.Arthritis Care Res (Hoboken) 2016;68(1):1-25.doi: 10.1002/acr.22783 [published Online First: 2015/11/08]

20. Pfizer.Salazopyrin tablets. Healthcare Professionals (SmPC) 2021 [updated 24/12/2021.2021:[Available from: https://www.medicines.org.uk/emc/product/3838/smpc#gref accessed 23/07/2023 2023.

21. Herrett E, Gallagher AM, Bhaskaran K, et al.Data Resource Profile: Clinical Practice Research Datalink (CPRD).International journal of epidemiology 2015;44(3):827–36.doi: 10.1093/ije/dyv098 [published Online First: 2015/06/08]

22. Wolf A, Dedman D, Campbell J, et al.Data resource profile: Clinical Practice Research Datalink (CPRD) Aurum.International journal of epidemiology 2019;48(6):1740–40g.doi: 10.1093/ije/dyz034 [published Online First: 2019/03/13]

23. Abhishek A, Doherty M, Kuo CF, et al.Rheumatoid arthritis is getting less frequent-results of a nationwide population-based cohort study.Rheumatology (Oxford) 2017;56(5):736–44.doi: 10.1093/rheumatology/kew468 [published Online First: 2017/01/09]

24. Nakafero G, Grainge MJ, Williams HC, et al.Risk stratified monitoring for methotrexate toxicity in immune mediated inflammatory diseases: prognostic model development and validation using primary care data from the UK.Bmj 2023;381:e074678.doi: 10.1136/bmj-2022-074678 [published Online First: 2023/05/31]

25. Nakafero G, Grainge MJ, Card T, et al.What is the incidence of methotrexate or leflunomide discontinuation related to cytopenia, liver enzyme elevation or kidney function decline? Rheumatology 2021;60(12):5785–94.doi: 10.1093/rheumatology/keab254

26. Khwaja A.KDIGO clinical practice guidelines for acute kidney injury.Nephron Clin Pract 2012;120(4):c179–84.doi: 10.1159/000339789 [published Online First: 2012/08/15]

27. Chalasani N, Björnsson E.Risk factors for idiosyncratic drug-induced liver injury.Gastroenterology 2010;138(7):2246–59.doi: 10.1053/j.gastro.2010.04.001 [published Online First: 2010/04/17]

28. Safy-Khan M, de Hair MJH, Welsing PMJ, et al.Current Smoking Negatively Affects the Response to Methotrexate in Rheumatoid Arthritis in a Dose-responsive Way, Independently of Concomitant Prednisone Use.J Rheumatol 2021;48(10):1504–07.doi: 10.3899/jrheum.200213 [published Online First: 2021/02/03]

29. Klotz U.Clinical Pharmacokinetics of Sulphasalazine, Its Metabolites and Other Prodrugs of 5-Aminosalicylic Acid.Clinical Pharmacokinetics 1985;10(4):285–302.doi: 10.2165/00003088-198510040-00001

30. Meijer B, Wilhelm AJ, Mulder CJJ, et al.Pharmacology of Thiopurine Therapy in Inflammatory Bowel Disease and Complete Blood Cell Count Outcomes: A 5-Year Database Study.Ther Drug Monit 2017;39(4):399–405.doi: 10.1097/ftd.0000000000000414 [published Online First: 2017/05/11]

31. Dirven L, Klarenbeek NB, van den Broek M, et al.Risk of alanine transferase (ALT) elevation in patients with rheumatoid arthritis treated with methotrexate in a DAS-steered strategy.Clin Rheumatol 2013;32(5):585–90.doi: 10.1007/s10067-012-2136-8 [published Online First: 2012/12/12]

32. Riley RD, Ensor J, Snell KIE, et al.Calculating the sample size required for developing a clinical prediction model.BMJ 2020;368:m441.doi: 10.1136/bmj.m441

33. Riley RD, Debray TPA, Collins GS, et al.Minimum sample size for external validation of a clinical prediction model with a binary outcome.Statistics in medicine 2021;40(19):4230–51.doi: 10.1002/sim.9025

34. Schafer JL.Multiple imputation: a primer.Stat Methods Med Res 1999;8(1):3–15.doi: 10.1177/096228029900800102 [published Online First: 1999/05/29]

35. Steyerberg EW. Clinical Prediction Models : A Practical Approach to Development, Validation, and Updating.Cham, SWITZERLAND: Springer International Publishing AG 2019.

36. Royston P, Altman DG.External validation of a Cox prognostic model: principles and methods.BMC Medical Research Methodology 2013;13(1):33.doi: 10.1186/1471-2288-13-33

37. Cox DR.Note on Grouping.Journal of the American Statistical Association 1957;52(280):543–47.doi: 10.1080/01621459.1957.10501411

38. Moons KG, Altman DG, Reitsma JB, et al.Transparent Reporting of a multivariable prediction model for Individual Prognosis or Diagnosis (TRIPOD): explanation and elaboration.Annals of internal medicine 2015;162(1):W1–73.doi: 10.7326/m14-0698 [published Online First: 2015/01/07]

39. Ricart E, Taylor WR, Loftus EV, et al.N-acetyltransferase 1 and 2 genotypes do not predict response or toxicity to treatment with mesalamine and sulfasalazine in patients with ulcerative colitis.Am J Gastroenterol 2002;97(7):1763–8.doi: 10.1111/j.1572-0241.2002.05838.x [published Online First: 2002/07/24]

40. Mielnik P, Sexton J, Fagerli KM, et al.Discontinuation rate of sulfasalazine, leflunomide and methotrexate due to adverse events in a real-life setting (NOR-DMARD).Rheumatol Adv Pract 2023;7(2):rkad053.doi: 10.1093/rap/rkad053 [published Online First: 2023/07/11]

41. Mori S, Arima N, Ito M, et al.Non-alcoholic steatohepatitis-like pattern in liver biopsy of rheumatoid arthritis patients with persistent transaminitis during low-dose methotrexate treatment.PLoS One 2018;13(8):e0203084.doi: 10.1371/journal.pone.0203084 [published Online First: 2018/08/25]

42. Nakafero G, Grainge MJ, Card T, et al.Development and validation of a prognostic model for leflunomide discontinuation with abnormal blood tests during long-term treatment: cohort study using data from the Clinical Practice Research Datalink Gold and Aurum.Rheumatology (Oxford) 2022;61(7):2783–91.doi: 10.1093/rheumatology/keab790 [published Online First: 2021/11/01]

43. Dirven L, Klarenbeek NB, van den Broek M, et al.Risk of alanine transferase (ALT) elevation in patients with rheumatoid arthritis treated with methotrexate in a DAS-steered strategy.Clinical rheumatology 2013;32(5):585–90.doi: 10.1007/s10067-012-2136-8

44. Cavalli M, Eriksson N, Sundbaum JK, et al.Genome-wide association study of liver enzyme elevation in an extended cohort of rheumatoid arthritis patients starting low-dose methotrexate.Pharmacogenomics 2022;23(15):813–20.doi: 10.2217/pgs-2022-0074

45. Sherbini AA, Gwinnutt JM, Hyrich KL, et al.Rates and predictors of methotrexate-related adverse events in patients with early rheumatoid arthritis: results from a nationwide UK study.*Rheumatology (Oxford*, England) 2022;61(10):3930–38.

46. Suzuki Y, Hirose T, Sugiyama N, et al.Post-marketing surveillance of high-dose methotrexate (>8 mg/week) in Japanese patients with rheumatoid arthritis: A post hoc sub-analysis of patients according to duration of prior methotrexate use.Modern Rheumatology 2021;31(3):575–86.

47. Verstappen SM, Bakker MF, Heurkens AH, et al.Adverse events and factors associated with toxicity in patients with early rheumatoid arthritis treated with methotrexate tight control therapy: the CAMERA study.Annals of the rheumatic diseases 2010;69(6):1044–48.doi: 10.1136/ard.2008.106617

48. Schmajuk G, Miao Y, Yazdany J, et al.Identification of risk factors for elevated transaminases in methotrexate users through an electronic health record.Arthritis Care and Research 2014;66(8):1159–66.doi: 10.1002/acr.22294

49. Hastings R, Ding T, Butt S, et al.Neutropenia in patients receiving anti-tumor necrosis factor therapy.Arthritis Care Res (Hoboken) 2010;62(6):764–9.doi: 10.1002/acr.20037 [published Online First: 2010/06/11]

