## Supplementary material for "Risk-stratified monitoring for sulfasalazine toxicity: prognostic model development and validation"

Figure S13: Calibration of a prognostic model for SSZ discontinuation with abnormal monitoring blood-test results at 5 years in the validation cohort: stratified according to age. 15

Figure S14: Calibration of a prognostic model for SSZ discontinuation with abnormal monitoring blood-test results at 5 years in the validation cohort: stratified according to disease type.......16

Supplementary Table 1: Individual patient's characteristics at the midpoint of each decile of risk.

| Decile | Age± (yr.) | Sex | BMI (kg/m$^2$) | Alcohol | Disease | DM | CKD | Immune-suppress. drug | Statin | Anti-epileptic | Paracet amol | BTA | Cumulative probability of outcome (%) |
|---|---|---|---|---|---|---|---|---|---|---|---|---|---|
| 1 | 25-30 | F | 35.2 | Moderate | PSA | No | No | MTX | No | No | No | No | 5.25 |
| 2 | 35-40 | M | 23.0 | Non-drinker | RA | No | No | No | No | No | No | No | 6.19 |
| 3 | 25-30 | F | 21.6 | Low | PSA | No | No | No | No | No | No | No | 6.72 |
| 4 | 55-60 | F | 29.8 | Low | RA | No | No | No | No | No | No | No | 7.25 |
| 5 | 55-60 | M | 24.0 | Non-drinker | RA | No | No | No | No | No | Yes | No | 7.80 |
| 6 | 70-75 | F | 27.9 | Low | RA | No | No | Aza/6-MP | No | Yes | Yes | No | 8.45 |
| 7 | 45-50 | F | 25.3 | Low | RA | No | No | MTX | No | No | No | No | 9.29 |
| 8 | 75-80 | M | 27.7 | Non-drinker | RA | Yes | No | No | Yes | No | Yes | No | 10.52 |
| 9 | 65-70 | M | 30.6 | Low | PSA | No | Yes | No | No | No | Yes | No | 13.90 |
| 10 | 55-60 | F | 33.8 | Non-drinker | RA | No | No | MTX | No | No | No | Yes | 19.04 |

Aza/6-MP: - Azathioprine/6-Mercaptopurine; BMI: - Body Mass Index; BTA: - Blood Test abnormalities within 6 months of first primary care sulfasalazine prescription; CKD: - Chronic Kidney Disease; Anti-epileptics: - carbamazepine / valproate; DM: - diabetes mellitus; F: - female; M: - male; MTX: - methotrexate; PsA: - psoriatic arthritis; RA: - Rheumatoid Arthritis. ± 5-year age band. Exact age not shown for anonymity.

Figure S1: Study population selection criteria for model development

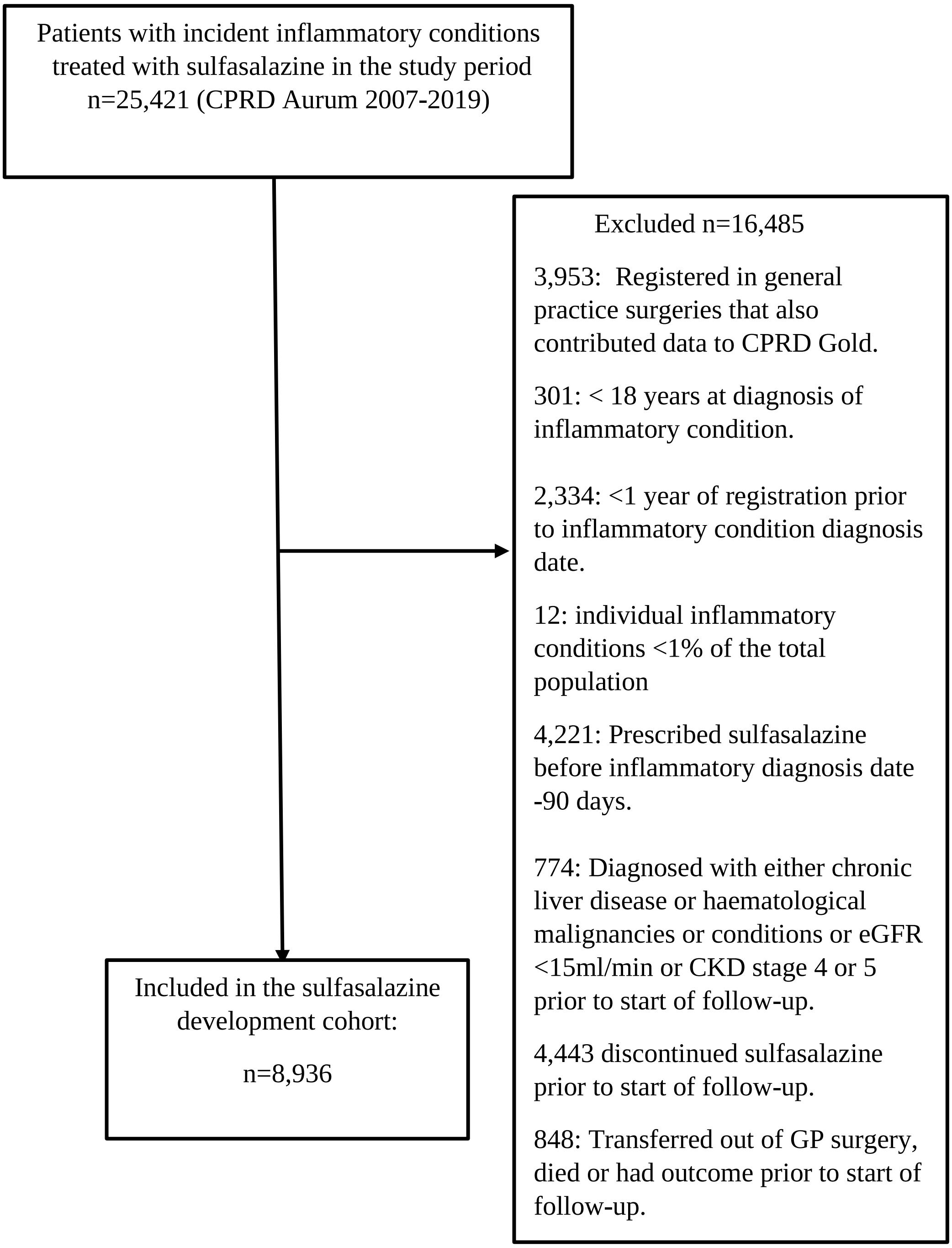

Figure S2: Model validation cohort: Study population selection criteria

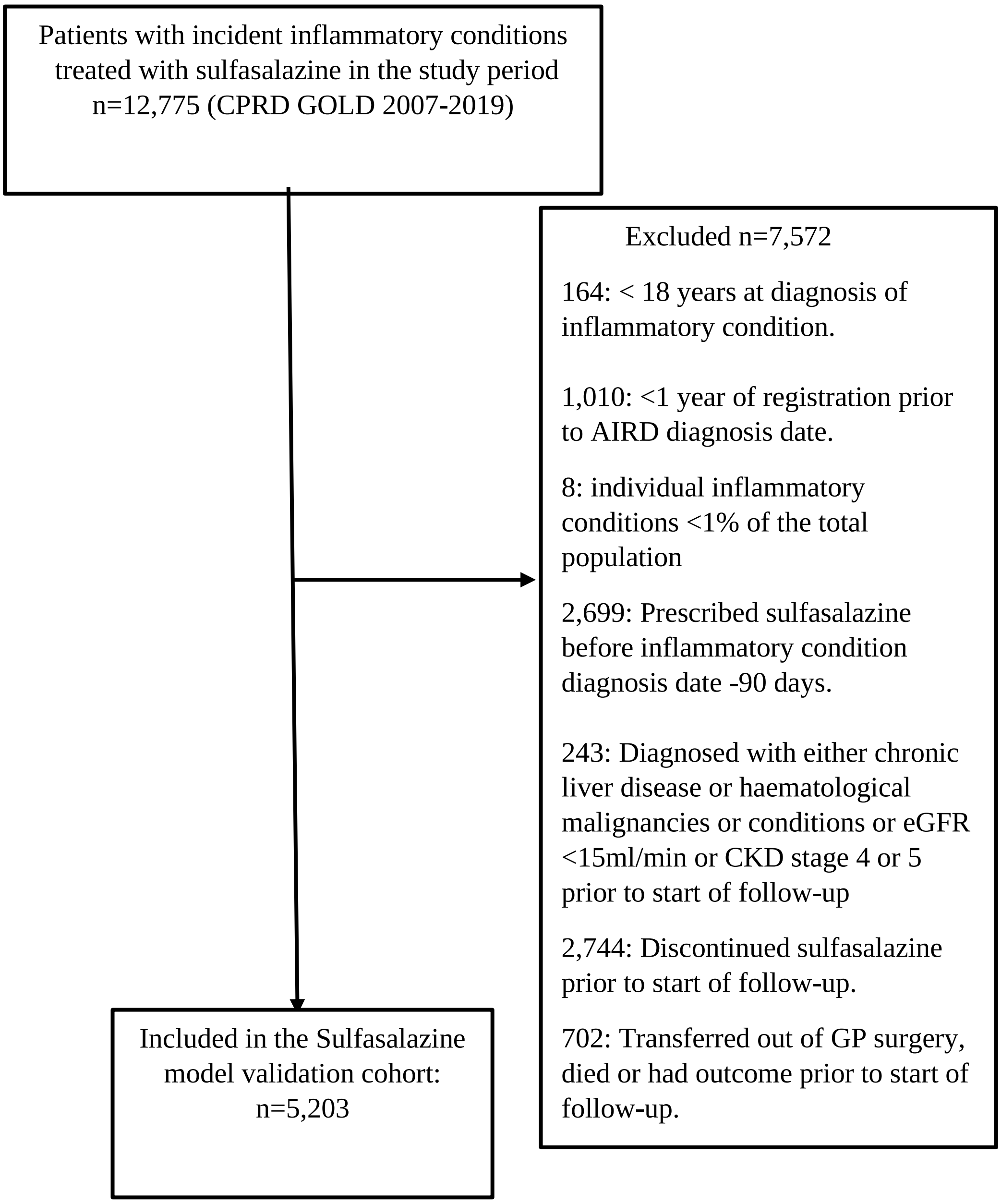

Figure S3: Occurrence of cytopenia, myelotoxicity and renal function decline in the cohort exposed to sulfasalazine

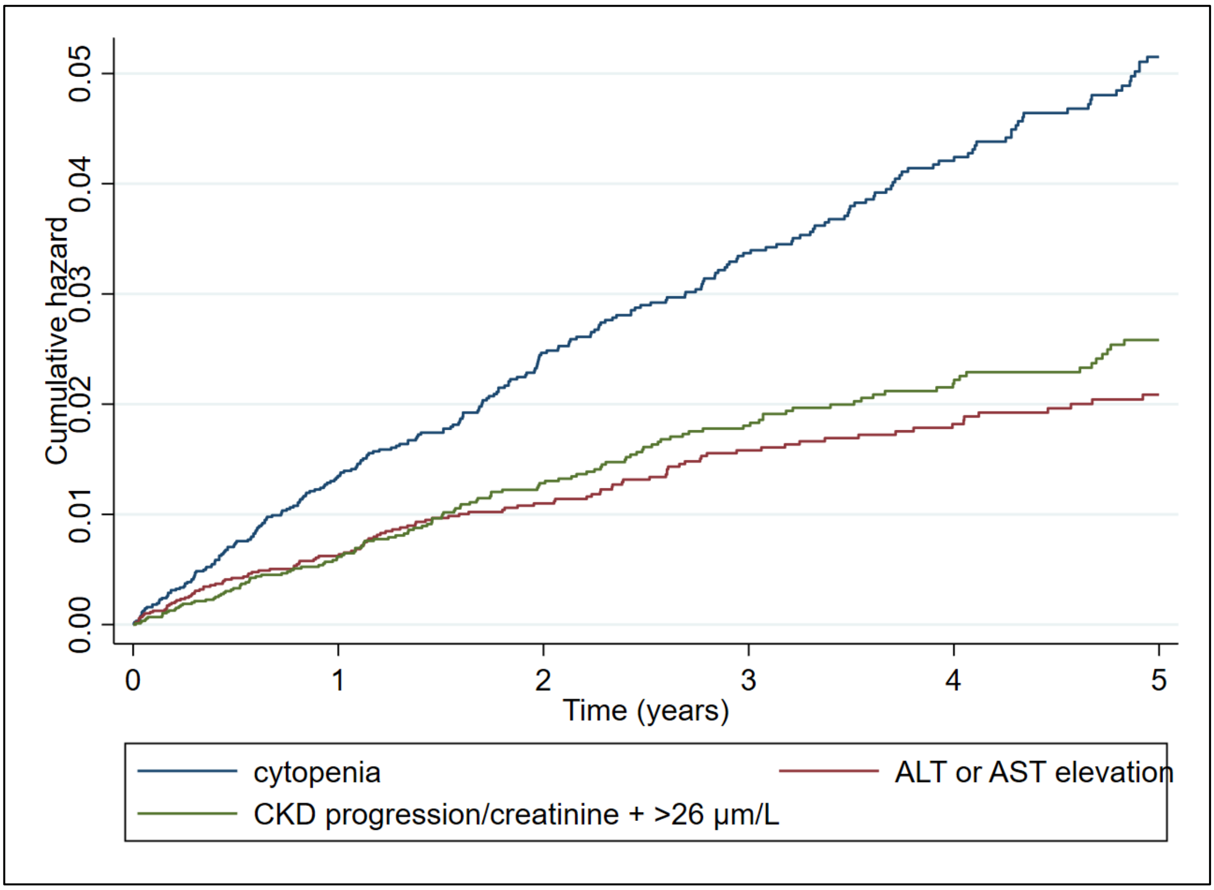

Figure S4: Occurrence of cytopenia, myelotoxicity and renal function decline in sulfasalazine prescribed cohort excluding patients prescribed either methotrexate, leflunomide, or thiopurines at cohort entry

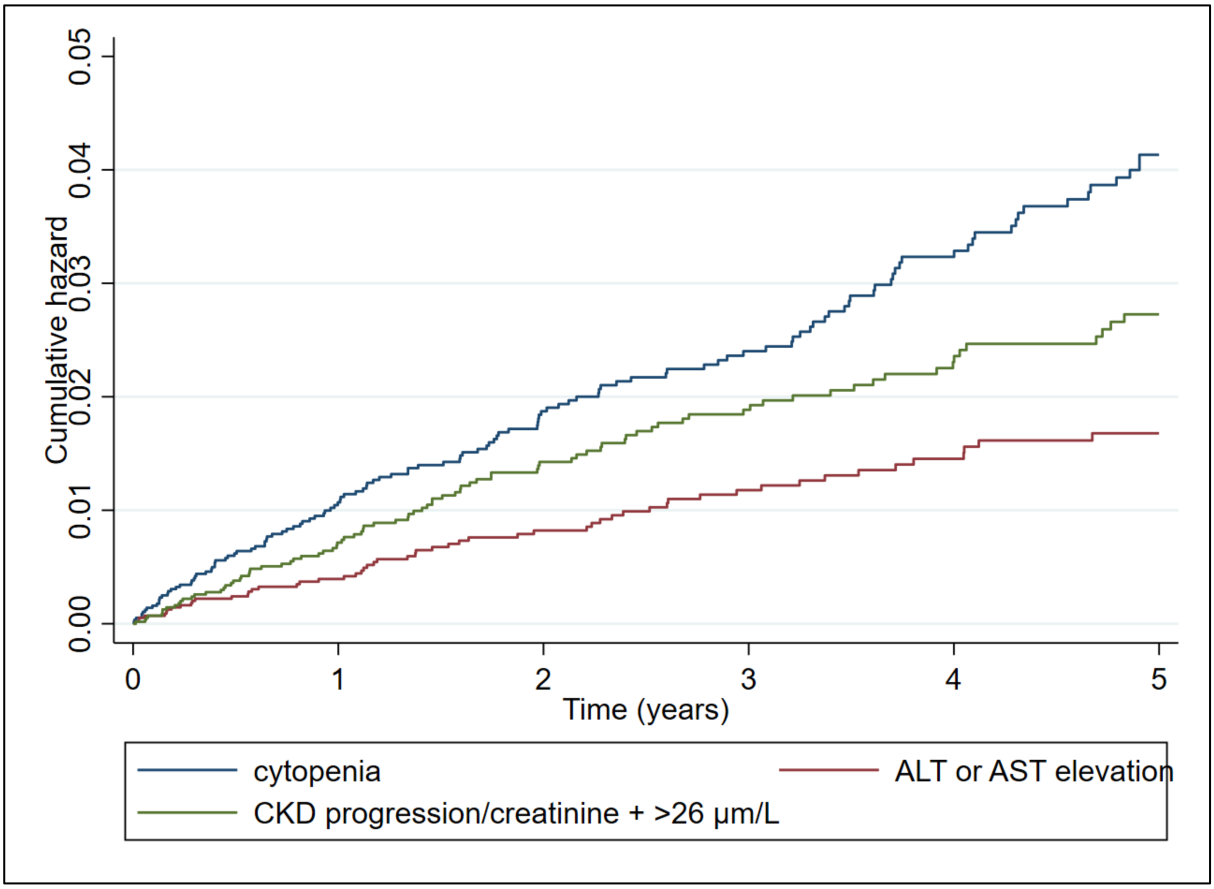

Figure S5: Distribution of predicted risk in the model derivation cohort at 5 years

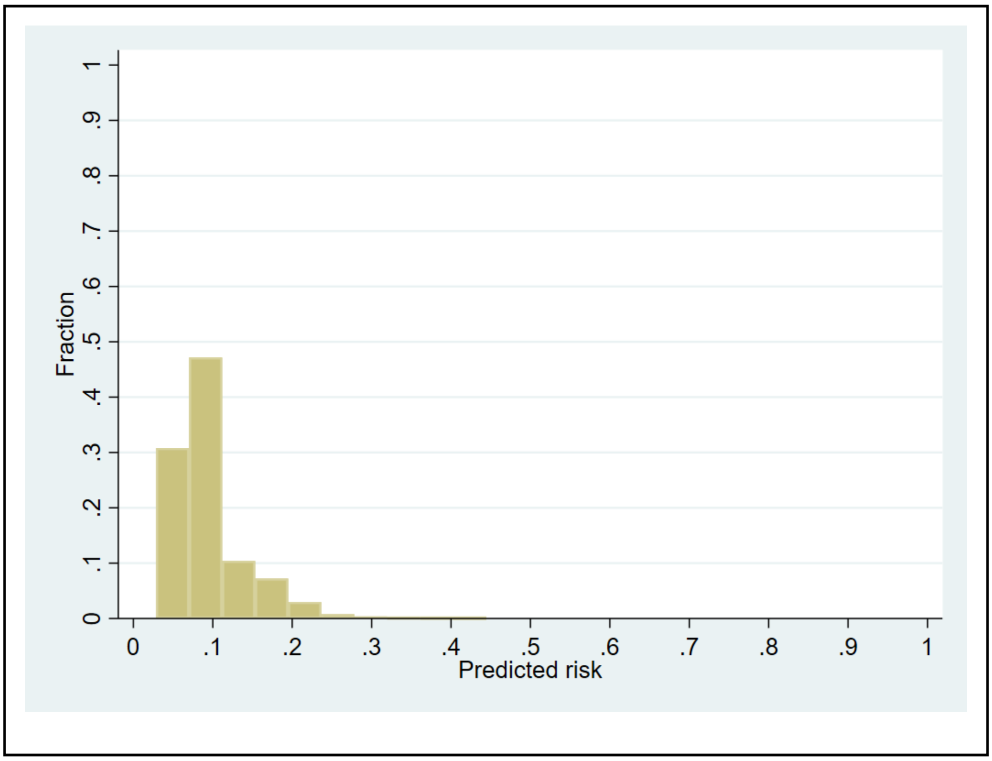

Figure S6: Calibration of a prognostic model for SSZ discontinuation with abnormal monitoring blood-test results at 5-years in the development cohort[1]

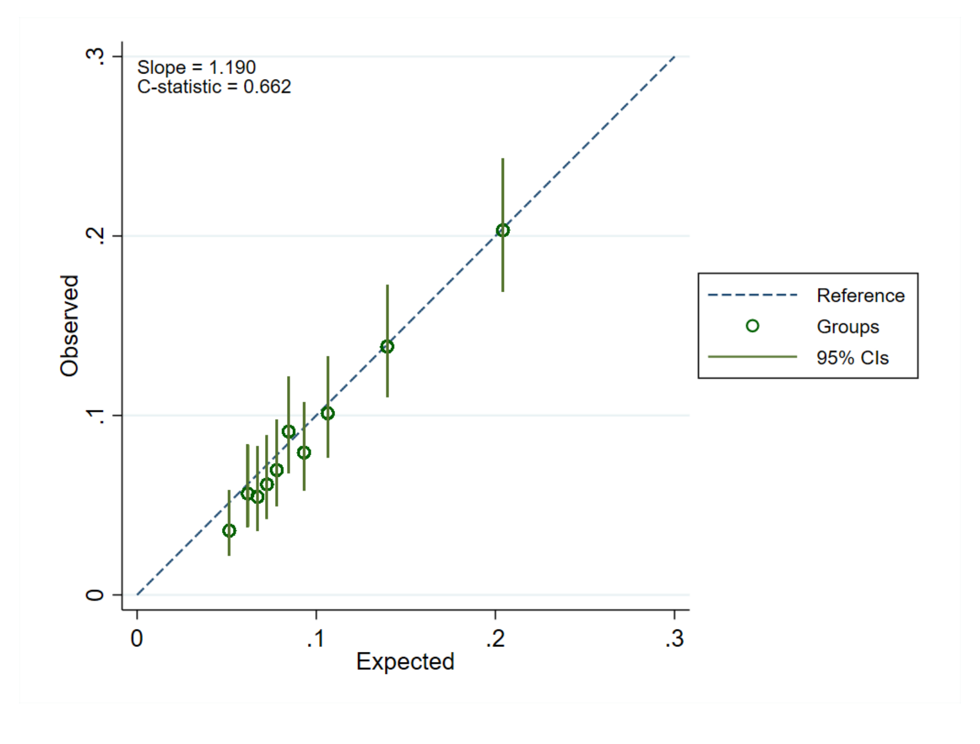

[1]Data from a single imputed dataset; So(t=5) 0.940

Figure S7: Calibration of a prognostic model for SSZ discontinuation with abnormal monitoring blood-test results at 5-years in the validation cohort[1]

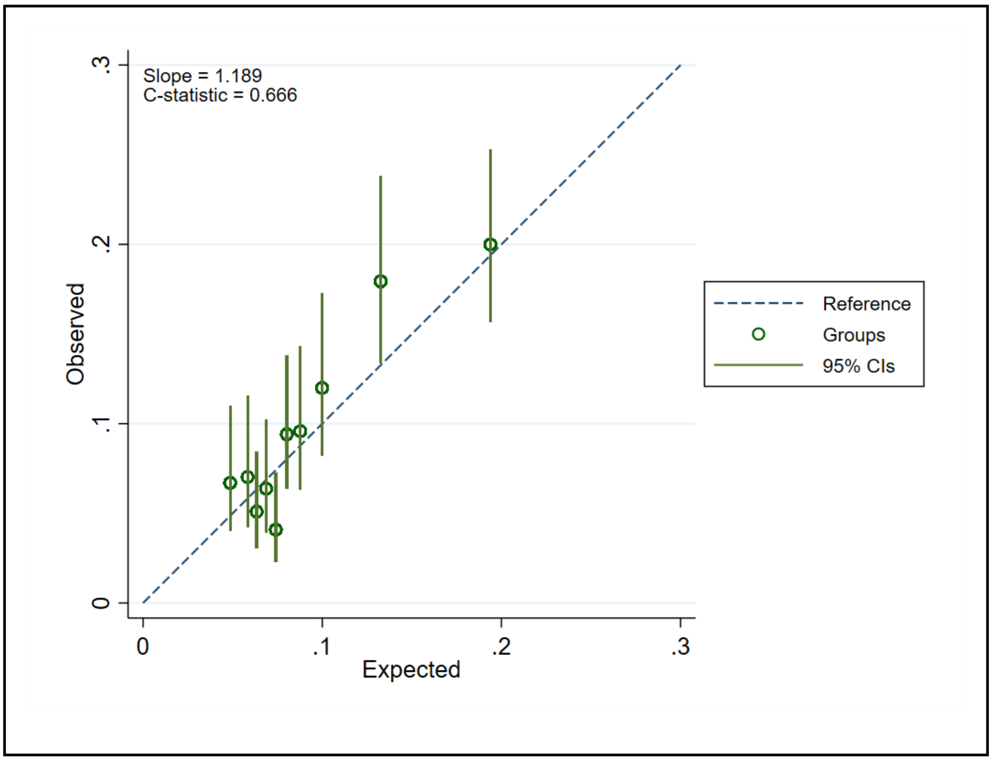

[1]Data from a single imputed dataset;  So(t=5) 0.940

Figure S8: Distribution of predicted risk in the model validation cohort at 5 years

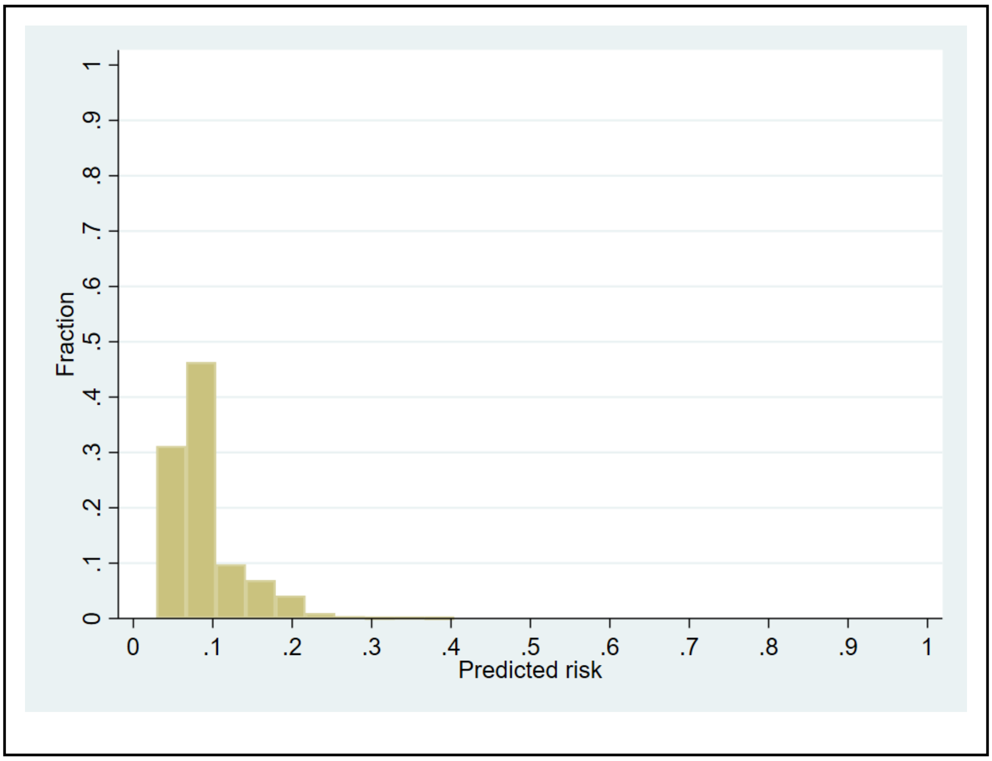

Figure S9: Calibration of a prognostic model for SSZ discontinuation with abnormal monitoring blood-test results at 1 year in the validation cohort[1]

A: Calibration plot

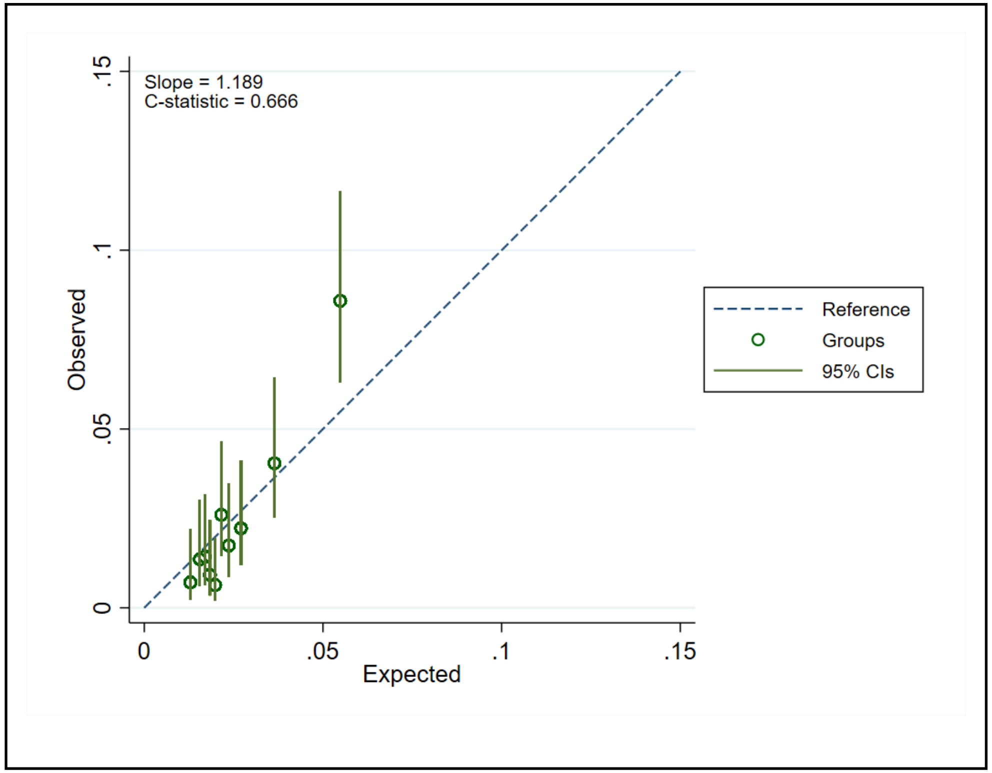

B: Smoothed calibration curve

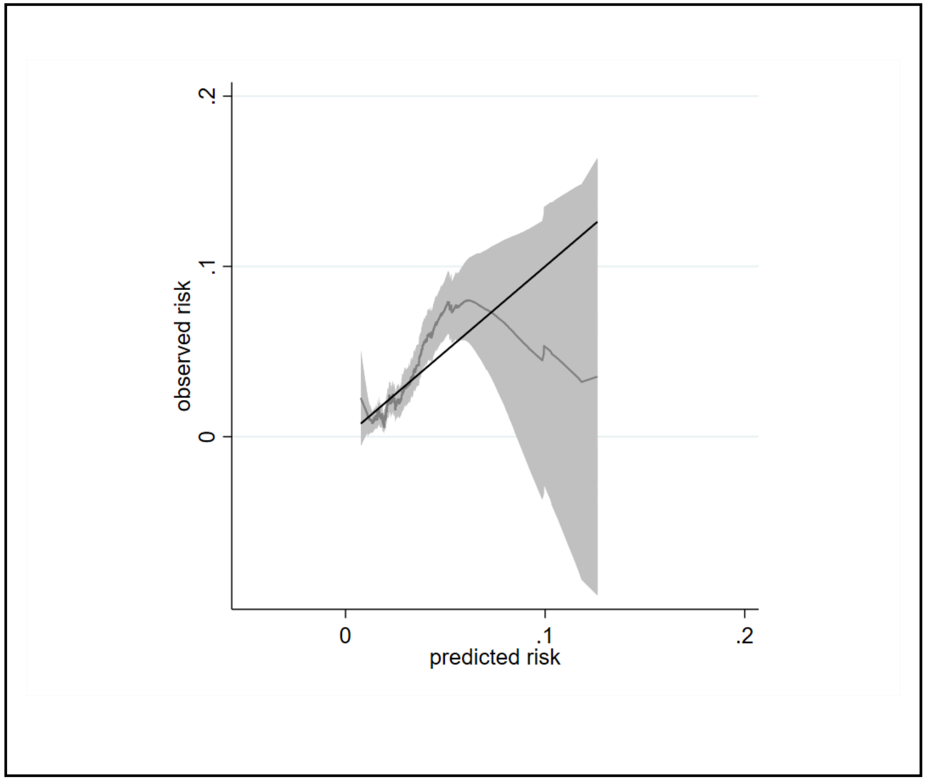

[1]Data from a single imputed dataset was used; $S_0(t_{=1})$ 0.984

Figure S10: Calibration of a prognostic model for SSZ discontinuation with abnormal monitoring blood-test results at 2 years in the validation cohort[1]

A. Calibration plot

B. Smoothed calibration curve

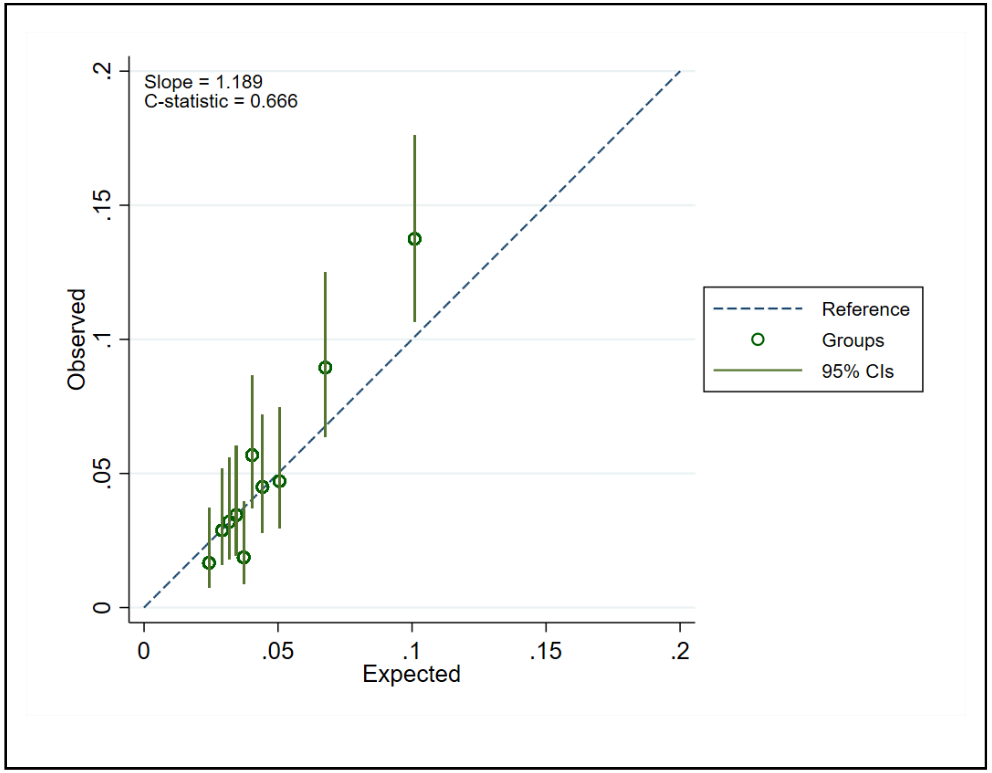

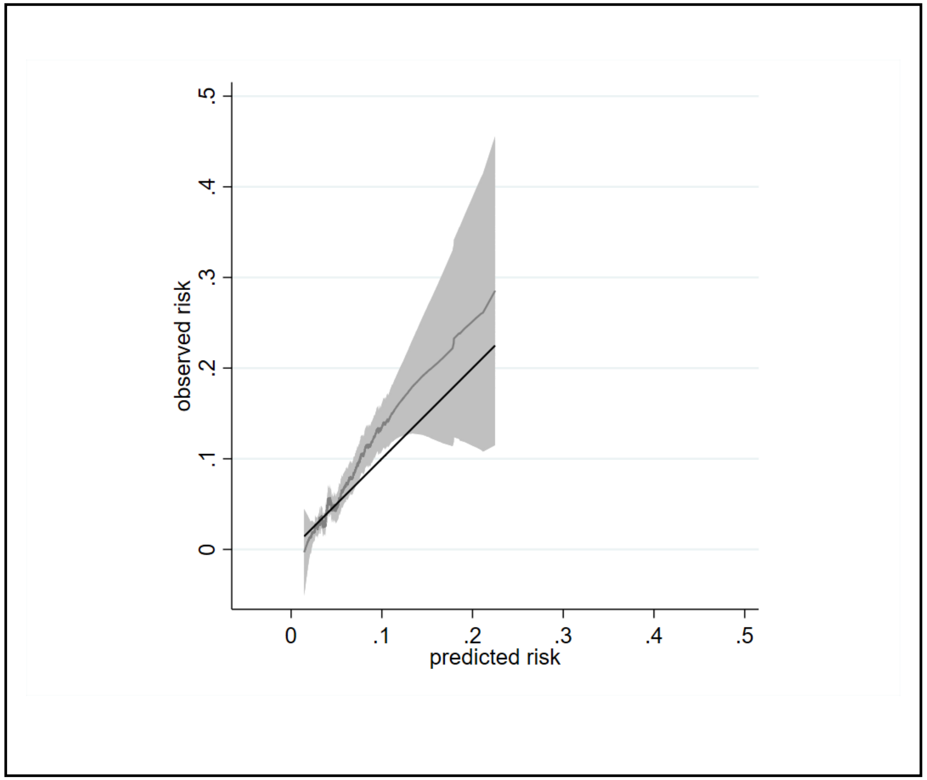

[1]Data from a single imputed dataset was used; $S_0(t_{=2})$ 0.970

Figure S11: Calibration of a prognostic model for SSZ discontinuation with abnormal monitoring blood-test results at 3 years in the validation cohort[1]

A. Calibration plot

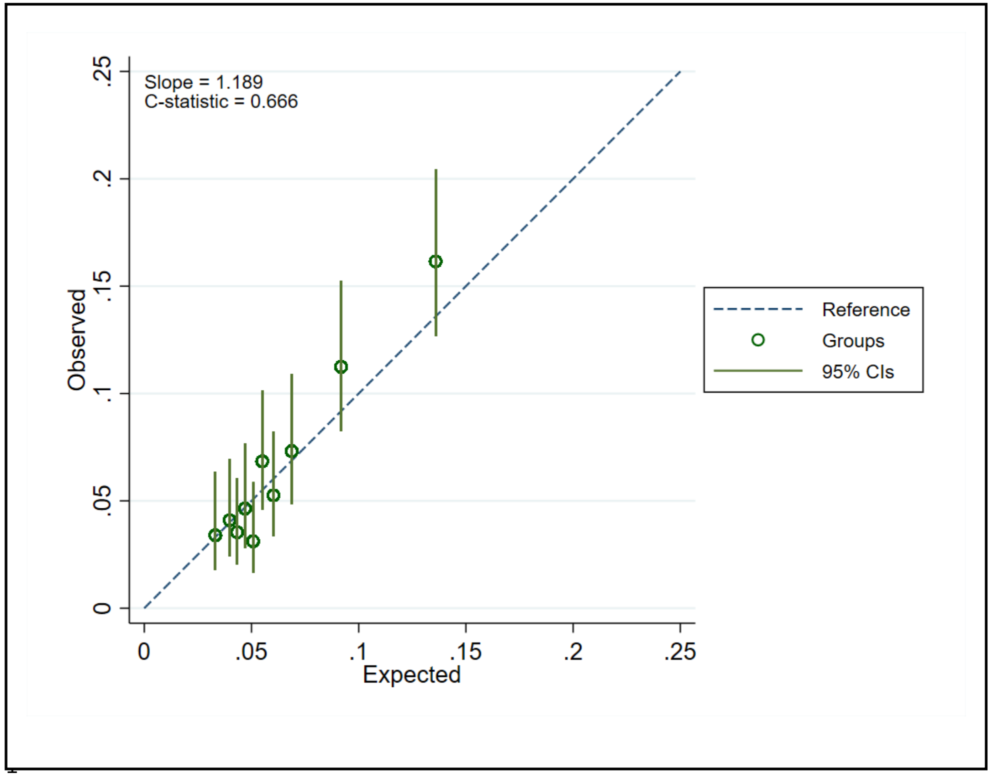

B. Smoothed calibration curve

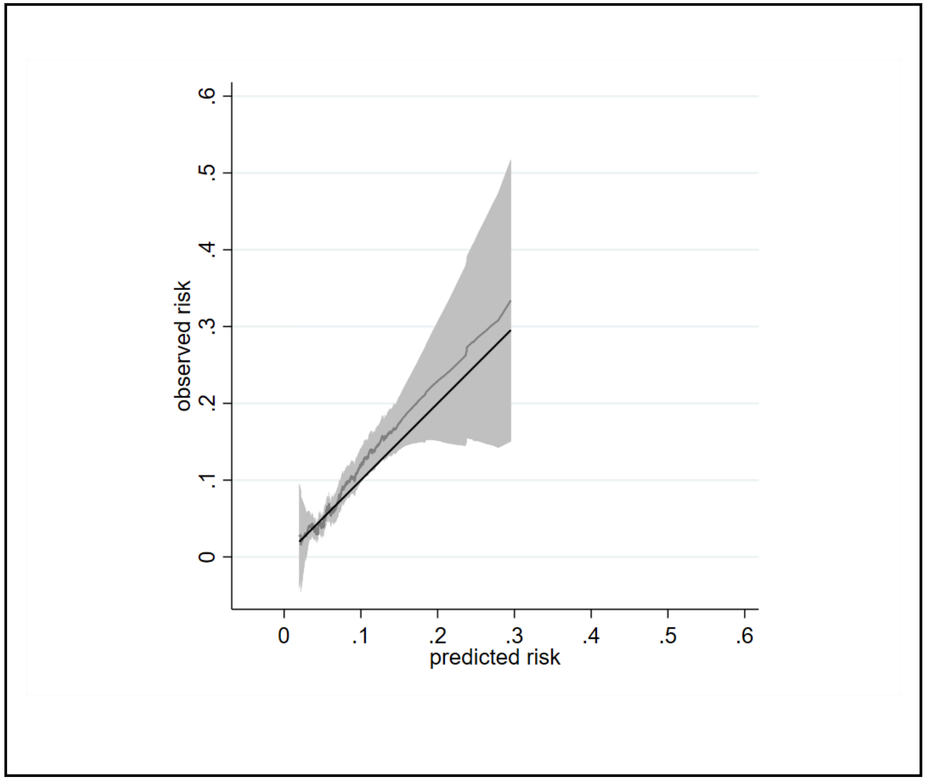

[1] Data from a single imputed dataset was used; $S_0(t_{=3})$ 0.959

Figure S12: Calibration of a prognostic model for SSZ discontinuation with abnormal monitoring blood-test results at 4 years in the validation cohort[1]

A. Calibration plot

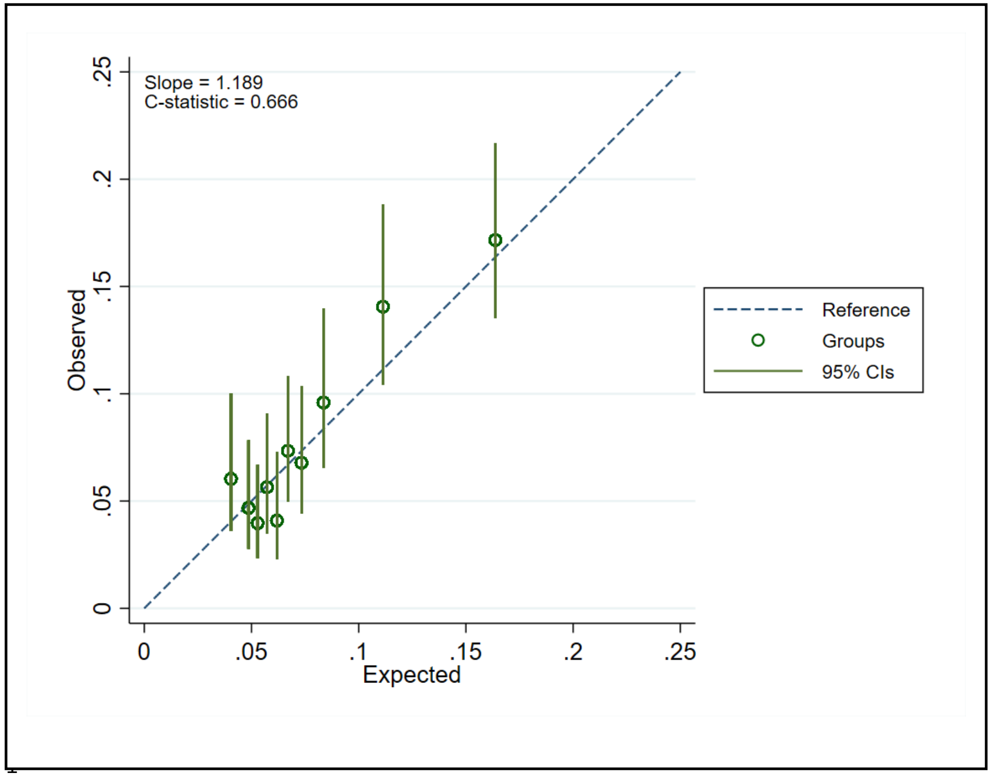

B. Smoothed calibration curve

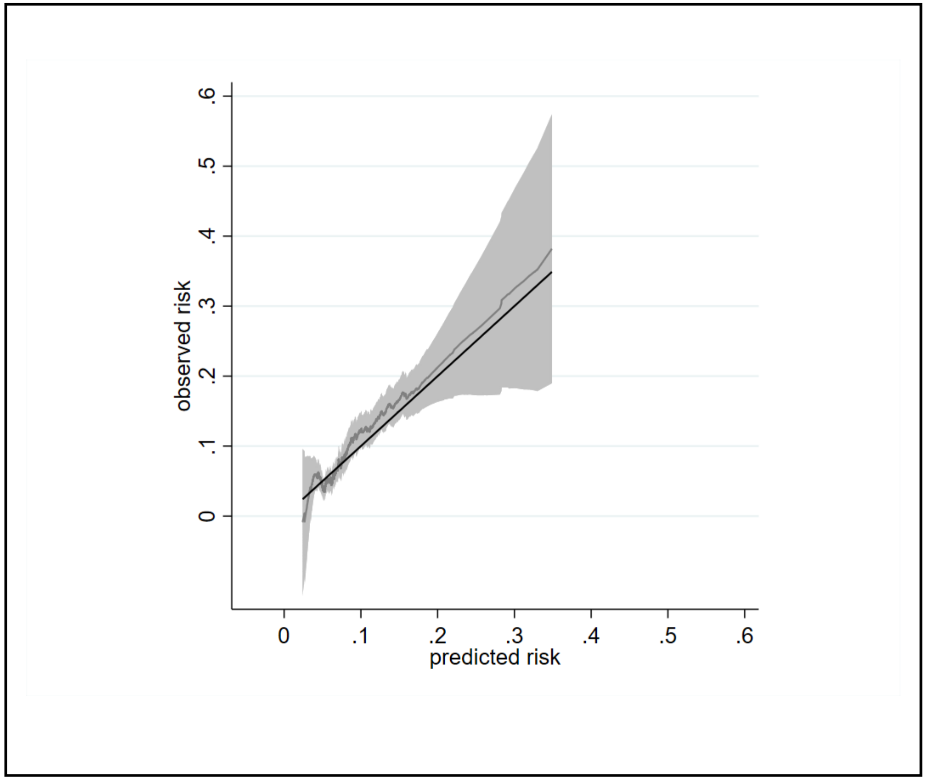

Data from a single imputed dataset was used; $S_0(t_{=4})$ 0.950

Figure S13: Calibration of a prognostic model for SSZ discontinuation with abnormal monitoring blood-test results at 5 years in the validation cohort: stratified according to age.

A. <60 years B. ≥ 60 years

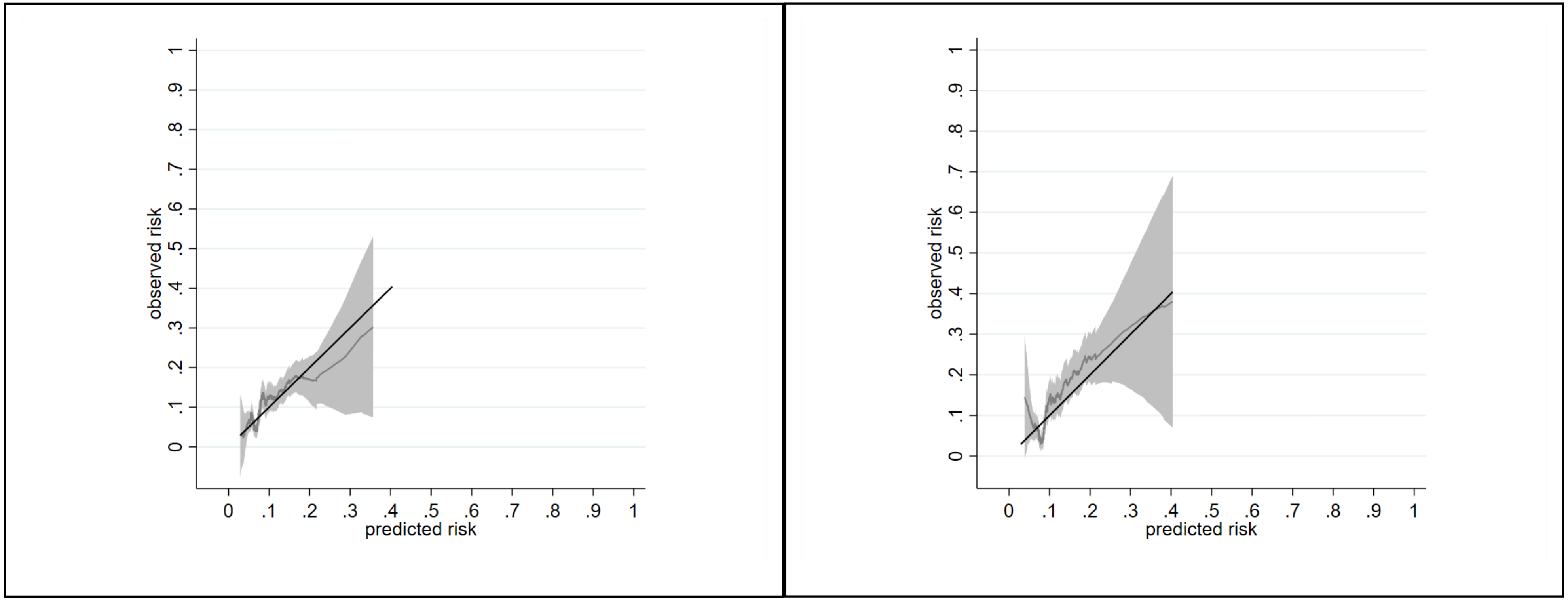

Figure S14: Calibration of a prognostic model for SSZ discontinuation with abnormal monitoring blood-test results at 5 years in the validation cohort: stratified according to disease type.

A: Rheumatoid Arthritis

B: other inflammatory conditions

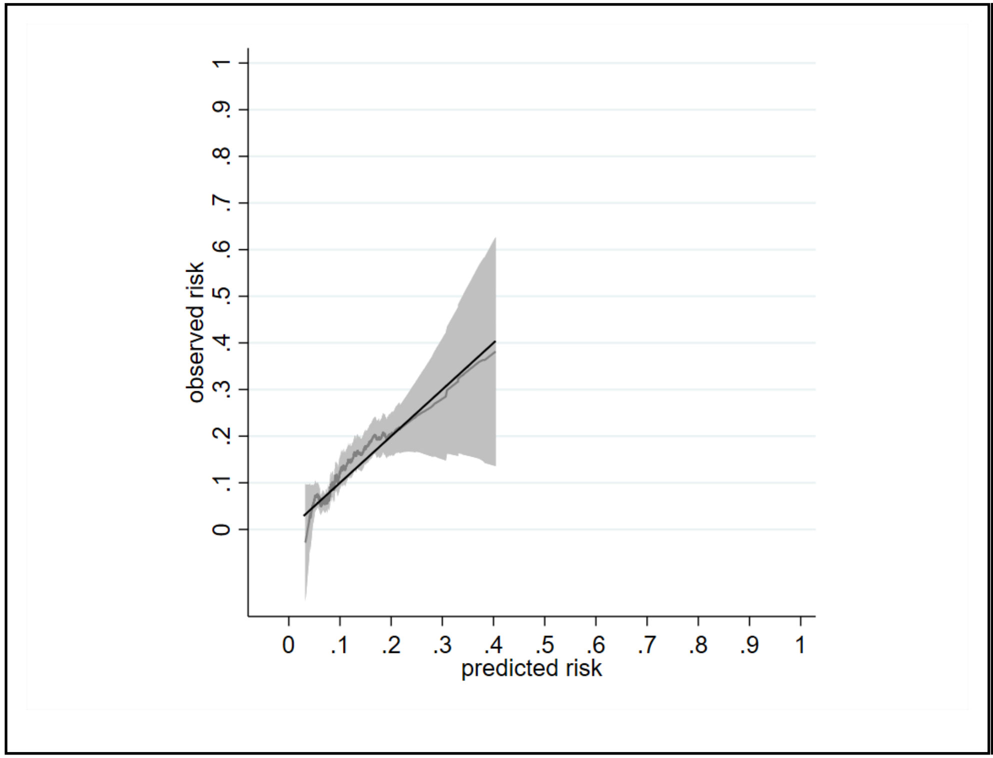

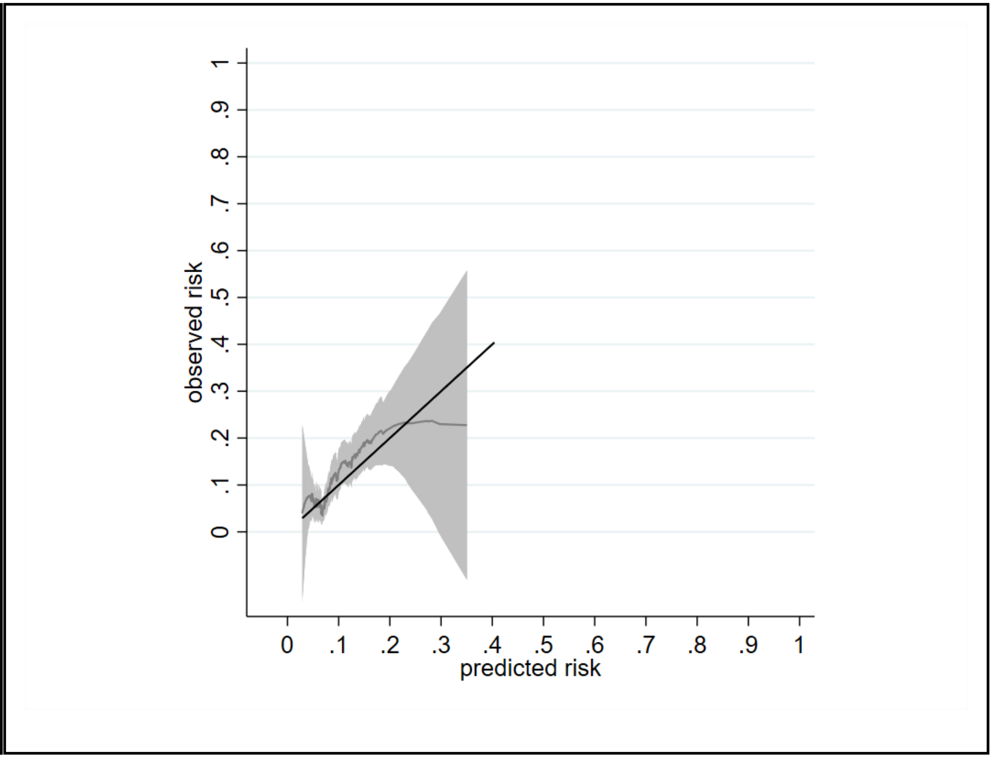
